# Decision-Making Under Stress, Biological Stress Responses, and Risk for Suicidal Behavior in Young Adults

**DOI:** 10.64898/2026.10.30.26366933

**Authors:** Nermin Toukhy, Meilin Jia-Richards, Eli Goodfriend, Violet Hecmanczsuk, Sarah Riston, Isabel Novacich, Nicolette Meade, Brian Thoma, Dara Sakolsky, Antoine Douaihy, Kehui Chen, David Brent, Susan Perlman, Anna Marsland, Nadine M. Melhem

**Affiliations:** Department of Psychiatry, University of Pittsburgh, School of Medicine, Pittsburgh, PA, USA; University of Pittsburgh Medical Center, Pittsburgh, PA, USA; Western Psychiatric Institute and Clinic of the University of Pittsburgh Medical Center, Pittsburgh, PA, USA; Department of Statistics, University of Pittsburgh, Pittsburgh, PA, USA; Department of Psychiatry, Washington University at St. Louis; Department of Psychology, University of Pittsburgh, Pittsburgh, PA, USA

**Keywords:** Decision-making, suicidal risk, biomarkers, stress, inflammation, hair cortisol

## Abstract

**Background:** Stress may influence decision-making processes relevant to suicidal behavior, yet few studies have examined risky decision-making and expected value (EV) sensitivity under stress longitudinally among young adults at elevated suicide risk.

**Methods:** Participants included 233 young adults (18–30 years) with recent suicide attempts (n=55), suicidal ideation (n=59), psychiatric controls (n=71), and healthy controls (n=48). Hair cortisol concentration, circulating inflammatory markers (CRP, IL-6, IL-10, IL-1ra, TNF-α), Stressful Cups Task, and clinical measures were assessed at baseline, 3-, 6-, and 12-month follow-ups.

**Results:** Participants with a history of suicide attempts showed greater reductions in risky choices under stress over time compared to healthy controls (β=−0.119, p<0.001). Greater reductions in EV sensitivity under stress relative to neutral conditions predicted subsequent suicidal behavior (OR=0.530, 95% CI: 0.328–0.856, p=0.009). Higher C-reactive protein (CRP) was associated with fewer risky choices under both neutral (β = −0.040, SE = 0.010, p < 0.001) and stress conditions (β = −0.131, SE = 0.031, p < 0.001), while higher interleukin-6 (IL-6) was associated with fewer risky choices under neutral conditions (β = −0.080, SE = 0.018, p < 0.001). Higher interleukin-10 (IL-10) was associated with greater reductions in risky choices under stress relative to neutral conditions over time (β = −0.068, SE = 0.018, p < 0.001). No significant associations were observed for hair cortisol concentrations (HCC).

**Conclusions:** Stress-related changes in risky decision-making and reduced EV sensitivity may contribute to vulnerability to suicidal behavior. These findings highlight the importance of assessing decision-making under stress and examining inflammatory processes as potential mechanisms underlying suicide risk.

## Introduction

Suicide is a major public health concern and remains one of the leading causes of death among young adults in the United States, ranking as the second leading cause of death for individuals between the ages of 20 and 34 ^1^. Stress plays a central role in suicidal thoughts and behaviors (STBs) ^2^ and has been shown to influence decision-making processes ^3^. In addition, biological stress responses, including cortisol and inflammatory markers, have been associated with risky decision-making ^4,5^ and suicidal behavior ^6^. Given that suicidal crises often occur in the context of acute stress, understanding the relationships between stress, biological stress responses, and decision-making may provide important insights into the mechanisms underlying suicidal behavior. However, few studies have examined these relationships among young adults at elevated risk for suicidal behavior, particularly using longitudinal designs.

A growing body of literature suggests that individuals with a history of STBs show altered decision making ^7,8^. Meta-analyses and systematic reviews indicate that suicide attempters (SA), compared to psychiatric controls (PC) and healthy controls (HC), tend to demonstrate riskier and more disadvantageous decision-making on laboratory-based paradigms such as the Iowa Gambling Task (IGT) and the Cambridge Gambling Task (CGT) ^8,9^. These studies found small-to-moderate effects indicating riskier decision-making among individuals with suicidal behavior, particularly those with a history of suicide attempts ^8,9^. Similarly, studies among adolescents have demonstrated impairments in risky decision-making associated with suicidal behaviors ^10^. Thus, decision-making processes could represent a promising cognitive marker of suicide risk.

Previous research on risky decision-making has distinguished between overall risk-taking and sensitivity to the expected value (EV) of available options ^11^. While overall risk-taking reflects the tendency to choose risky over safe options, EV sensitivity reflects the extent to which individuals adjust their choices according to the relative expected values of available options, favoring risk when advantageous and avoiding it when disadvantageous ^11^. Commonly used paradigms such as the Iowa Gambling Task (IGT) do not clearly distinguish between advantageous and disadvantageous risky choices, making it difficult to determine whether risky decision-making reflects generalized risk-taking or sensitivity to EV differences. Few studies examined value sensitivity in young adults at risk for suicidal behaviors. One study found that depressed suicide attempters were less able to discriminate between closely valued alternatives than individuals with suicidal ideation, non-suicidal depressed patients, and healthy controls ^12^. In contrast, individuals with major depression showed value sensitivity similar to healthy controls, suggesting that altered value sensitivity may not characterize depression generally ^13^. Thus, further work is needed to examine EV sensitivity among young adults at risk for suicidal behavior and determine whether impaired EV sensitivity is associated with suicide risk.

Stress may further influence decision-making processes associated with suicidal behavior. Previous studies have demonstrated that acute stress can shift decision-making toward more reward-driven or riskier patterns in both clinical and non-clinical populations ^3,14,15^. More specifically, one study demonstrated that stress exposure amplified risk-taking decisions among individuals with a history of suicidal behaviors relative to healthy controls ^14^. However, most prior studies examining decision-making among suicidal individuals have not incorporated stress into decision-making paradigms. Furthermore, the majority of studies have been cross-sectional, limiting the ability to determine whether differences in decision-making represent stable characteristics or dynamically change over time ^8,9^. Therefore, examining decision-making under stress longitudinally may provide important insights into cognitive processes associated with suicidal behavior.

Biological stress responses may also play a role in decision-making processes relevant to suicidal behavior. Studies in healthy adults indicate that exogenously administered cortisol can influence risky decision-making, particularly in reward-salient contexts ^4,16^. In addition, studies using stress-induction tasks indicate that individual differences in cortisol reactivity are associated with risky decision-making, such that smaller cortisol elevations are associated with riskier economic and social decisions ^17^, while elevated cortisol during stress has been linked to poorer performance on gambling tasks ^18^. A study among healthy adolescents further demonstrated that lower hair cortisol concentrations were associated with greater risky decision-making ^19^. Emerging evidence also suggests that systemic inflammation is associated with impaired decision-making processes in adults with depression and suicidal behaviors ^6^. Higher inflammatory markers, including IL-6 and TNF-α, have been associated with more present-focused and impulsive decision-making in non-clinical samples ^5^. Taken together, these findings suggest that biological stress responses may be associated with decision-making processes, although their relationships with risky decision-making and EV sensitivity among individuals at elevated risk for suicidal behavior remain understudied.

In this study, we examine the relationships of risky decision-making and EV sensitivity under neutral and stress conditions with suicide risk and biological markers of stress and inflammation, both cross-sectionally and longitudinally. We also examine whether risky decision-making and EV sensitivity prospectively predict subsequent suicide attempts and suicidal behaviors. We hypothesize that young adults with a recent suicide attempt or suicidal ideation will demonstrate greater risky decision-making and lower EV sensitivity compared to young adults with psychiatric diagnoses but no history of suicidal thoughts or behaviors and healthy controls, particularly under stress, both at baseline and over time; (2) greater risky decision-making and lower EV sensitivity, particularly under stress, will prospectively predict subsequent suicide attempts and suicidal behaviors; and (3) reduced hair cortisol concentration and high inflammation will be associated with risky decision-making and EV sensitivity, particularly under stress, both cross-sectionally and longitudinally.

## Method

### Participants

Psychiatric inpatients and outpatients 18 – 30 years old were recruited through the Western Psychiatric Hospital at the University of Pittsburgh Medical Center (UPMC). We recruited patients admitted for a suicide attempt (SA), participants with active suicidal ideation (SI), and psychiatric controls with no current suicidal ideation or behavior (PC). Healthy controls (HC) with no history of psychiatric disorders or suicidal ideation and behavior were recruited through a university participant registry.

Participants were excluded if they had a chronic condition affecting immune or endocrine pathways, were pregnant, or taking prescribed oral corticosteroids or other medications that affect immune and neuroendocrine functioning in the past 12 months, and if they had an infection or were on antibiotics in the past two weeks.

Assessments were conducted at baseline and at 3-, 6-, and 12-month follow-ups. The study was approved by the University of Pittsburgh Institutional Review Board and all participants provided their informed consent to participate in the study. We recruited a total of 515 participants of which 397 (77.1%) completed a baseline assessment. The analytic sample included 233 participants who completed our decision-making task, the Stressful Cups Task, at baseline,. The final sample consisted of 55 participants in the SA, 59 in the SI, 71 in the PC, and 48 in the HC groups.

The sample was 39% male, 71% White, with a mean age of 24.59 years (Standard Deviation or SD = 3.70). **Supplementary Table 1** compares participants included in the analyses (N=233) who completed the decision-making task to those who did not complete (N=164) on sociodemographic and clinical characteristics. Participants included in the analysis were more likely to be in the HC group and less likely to be LGBTQ compared to those excluded (**Supplementary Table 1**). In addition, those who completed the task had lower childhood trauma, affective lability, apathy, hopelessness, impulsivity, anxiety, insomnia, depressive symptoms, somatic symptoms, perceived stress and post-traumatic stress compared to those who did not complete the task (**Supplementary Table 1**).

Of the sample included in our analyses (N=233), 99 (42.5%) were lost to follow-up and N=134 (57.5%) completed at least one follow-up. **Supplementary Table 2** presents comparisons between those who were retained (had at least one follow-up completed) and those lost to follow-up (had only baseline data) on sociodemographic and clinical characteristics. Compared to those who were retained, those lost to follow-up were more likely to be in the SA group and less likely to be in the HC group, were more likely to be male, and had lower socioeconomic status and IQ. In addition, those lost to follow-up were more likely to have mood and substance use disorders and were more likely to have a lifetime history of suicidal behavior compared to those retained. Similarly, those who were lost to follow-up showed higher levels of clinical symptoms (affective lability, apathy, anxiety, depression etc.) (**Supplementary Table 2)**. Given the high attrition rate and significant differences in sociodemographic and clinical characteristics between participants retained and those lost to follow-up, inverse probability weighting (IPW) was applied in all longitudinal analyses to account for differential attrition and reduce potential attrition-related bias.

## Materials

### Sociodemographic and Psychiatric Characteristics

Participants self-reported demographic information (e.g., age, sex at birth, race) and completed the Hollingshed scale ^20^ as an estimate of socioeconomic status (SES). The Hollingshed scale calculates SES based on education level and income, with higher numbers indicating higher SES. Diagnoses were assessed using the Structured Clinical Interview for DSM-5 ^21^. After each interview, clinical interviewers met with a licensed psychiatrist (DS) to confirm diagnoses. Electronic health records were also reviewed for psychiatric history.

### Suicidal Thoughts and Behaviors

Current and past suicidal thoughts and behaviors were assessed using the Columbia Suicide Severity Rating Scale (C-SSRS) ^22^. Actual suicide attempt (SA) and suicidal behaviors including actual, aborted, interrupted, preparatory behavior) (SB) based on the C-SSRS and emergency referrals conducted during the study.

### Intelligence Quoffiecient

All participants completed the Vocabulary and Matrix Reasoning tests from the Wechsler Abbreviated Scale of Intelligence 2nd Edition (WASI-II) ^23^ to estimate participants’ IQ.

### The Stressful Cups Task

We used a modified version of the Cups task^24^ developed by S.P. to include a stress condition, The Stressful Cups Task. For each trial, participants are presented with a “safe” cup with a fixed monetary reward on one side of the screen and 2 – 5 “risky” cups that either gave no reward or a higher reward than the safe cup. The probability of winning a large reward in the high-risk condition varied between .2, .33, and .5. Trial types were classified as either “risk advantageous” where the expected value of risky choice was higher than the safe choice, “risk disadvantageous” where the expected value of the risky choice was lower than the safe choice, or “equivalent expected value” where expected values for both choices were equal. To assess decision-making under stress, the original cups task was modified by adding a stress condition where a loud aversive noise was played during the trial. The noise was a high-frequency white noise played through headphones for 12 of the 45 total trials.

Our primary outcomes on the Stressful Cups Task were the proportion of risky choices under neutral and stress conditions and their difference (stress minus neutral), and EV sensitivity under neutral and stress conditions and their difference (stress minus neutral). The proportion of risky choices was calculated as the proportion of trials in which participants selected the risky rather than the safe option across all trial types. EV sensitivity was calculated as the difference between the proportion of risky choices on risk-advantageous and risk-disadvantageous trials, consistent with the operationalization used in previous studies ^25,26^, with higher values indicating greater sensitivity to the relative expected values of the available options.

### Biological markers

#### Inflammatory markers

blood samples (10 ml) were collected during baseline and at 3, 6 and 12 month follow-ups to test for plasma levels of C-reactive protein (CRP), Interleukin-10 (IL-10), Interleukin-6 (IL-6), Interleukin-1 receptor antagonist (IL-1ra) and Tumor necrosis factor alpha (TNF-α). Plasma was collected from a blood draw. Samples for the assessment of cytokine levels were centrifuged for 15 min, and then immediately stored at −80 °C. Plasma levels of IL-6, TNFα, IL-1ra, and IL-10 were measured with Simple Plex assays using the antibody-based Ella™ system (ProteinSimple, Biotechne) per the manufacturer’s instructions. This system employs a solid-phase sandwich immunoassay run in triplicate. Levels of IL-6 and TNFα assessed using ELLA correlate highly with those assessed by ELISA (p’s > 0.97) ^27^. The average intra-assay coefficient of variations (CV) was 5.08%, 2.30%, 2.50%, and 6.57% for IL-6, TNFα, IL-1ra, and IL-10, respectively. High Sensitivity CRP was assayed per the manufacturer’s instructions using CRP Latex reagent (Beckman Coulter) on an AU5800 Series Clinical Chemistry Analyzer (Beckman Coulter, Inc., Brea, California).

#### Hair Cortisol Concentration (HCC)

was assessed by collecting approximately 50 mg of hair from 3cm closest to the scalp, at baseline and at 3, 6 and 12 month follow-ups. HCC collected from 3cm of hair provides a measure of cortisol levels over the past 3 months. This reflects cumulative cortisol secretion over this period of time, rather than momentary or acute fluctuations. As cortisol is incorporated into the hair during growth, HCC offers a stable, retrospective index of chronic HPA-axis activity that is less influenced by short-term stressors, diurnal variation, or situational reactivity ^28^. Hair samples were assayed using a highly sensitive enzyme immunoassay following methods described previously ^28^. All samples were analyzed in duplicate, with an average inter-assay CV of 4.74% and an intra-assay CV below 10% ^29,30^

#### BMI

was also assessed using participants’ index height and weight.

### Clinical Symptom Measures

Participants completed a battery of self-report measures assessing clinical symptoms associated with suicidality. To assess anxiety, depression, hopelessness, apathy, and anhedonia, we used the Generalized Anxiety Disorder-7 scale ^31^, Patient Health Questionnaire ^32^, Beck Hopelessness Scale ^33^, Apathy Evaluation Scale ^34^, and the Snaith-Hamilton Pleasure Scale ^35^. Aggression, perceived stress, symptoms of PTSD, impulsivity, and affect lability were measured using the Buss-Perry Aggression Questionnaire ^36^, Perceived Stress Scale ^37^, Post-traumatic Stress Disorder (PTSD) Checklist for the Diagnostic and Statistical Manual of Mental Disorders—5^th^ Edition ^38^, Barratt Impulsivity Scale ^39^, and the Affective Lability Scale ^40^. Lastly, we used the Drug Use Screening Inventory ^41^ and the PhenX Toolkit insomnia rating scale ^42^ to assess substance use and insomnia, respectively.

### Risk and Protective Factors

We assessed childhood trauma and social support as they are related to suicide risk ^43,44^. The Childhood Trauma Questionnaire (CTQ) ^45^ was used to assess childhood trauma, and the Multidimensional Scale of Perceived Social Support was used to assess social support ^46^.

### Statistical Analyses

All analyses were conducted using R version 4.5.1 ^47^. Given the number of clinical symptom measures used in the study, we conducted an exploratory factor analysis with varimax rotation to reduce the baseline data. A four-factor solution best fit the data statistically and clinically, which represented psychological distress, anhedonia, affect lability, and substance use (**Supplementary Table 3**). For follow-up timepoints, we used the factor structure identified at baseline and conducted confirmatory factor analyses for each follow-up timepoint. Measurement invariance was examined to confirm that the factor structure was consistent across timepoints, which suggested invariance indicating that the same 4-factor structure can be used in longitudinal analyses (**Supplementary Tables 4**)

Analyses of Stressful Cups Task performance were conducted separately for our six outcomes. Cross-sectional group differences (SA, SI, PC, HC) at baseline were examined using linear regression models. All models adjusted for IQ to account for potential differences in general cognitive ability, consistent with previous research using the Cups Task ^48^. Pairwise group comparisons were adjusted using Tukey’s procedure within each outcome, followed by a Bonferroni adjustment across the six outcomes (adjusted significance threshold: p < 0.0083).

Covariates were introduced hierarchically, including sociodemographic characteristics, clinical symptom factors, and risk and protective factors. Models were reduced at each step using backward stepwise regression.

We then examined changes in the Stressful Cups Task performance over time and whether there was a differential change over time by group. Linear mixed-effects models were conducted including group, time, and group by time interactions, with IQ retained in all models. Tukey and Bonferroni adjustments were applied as described above. Given the high attrition rate and significant clinical differences between retained and non-retained participants, longitudinal analyses were adjusted for attrition using inverse probability weighting (IPW)^49,50^. Retention probabilities were estimated using logistic regression including variables associated with retention (**Supplementary Table 2**), and stabilized weights were calculated using predicted retention probabilities truncated at 0.05. We conducted sensitivity analyses comparing models with and without IPW.

To examine the relationships between the biological measures and the six outcomes from the Stressful Cups Task, we first examined univariate associations at baseline between the biological measures and each outcome and corrected for multiple comparisons using the Benjamini–Yekutieli false discovery rate procedure. Significant associations at baseline were subsequently examined in adjusted models using the hierarchical covariate-selection strategy described above. Group, IQ and BMI were included in these models regardless of statistical significance because of their relevance to Stressful Cups Task performance and biological measures. We also examined whether the Stressful Cups Task performance changed over time with changes in biological measures. We used linear mixed-effects models including the main effect of each biological measure, time, and the biological measure by time interaction and adjusted for attrition using IPW. Sensitivity analyses were conducted for these models without IPW adjustment.

Finally, we examined whether the Stressful Cups Task performance predicted prospective actual suicide attempt (SA) and suicidal behavior (SB) events. We used logistic regression to examine whether baseline performance predicted events and Cox regression models to examine time to onset of SA and SB. Models were first examined without clinical covariates and subsequently adjusted for relevant clinical covariates. These prospective analyses additionally controlled for diagnosis and inpatient versus outpatient treatment status. HC participants were excluded from these analyses because no suicidal events were expected or occurred in this group.

## Results

### Sample Characteristics

**Table 1** presents the comparisons of SA, SI, PC, and HC on sociodemographic and clinical characteristics. Groups did not differ in terms of age or sex. Participants in the SA group had a lower proportion of White participants (59%, n = 32) compared to the SI (81%, n = 48) group. The SA (39%, n=19), SI (37%, n=20), and PC (34%, n=22) group had higher proportion of LGBTQ-identifying participants compared to HC group (8.5%, n = 4).

**Table 1.**
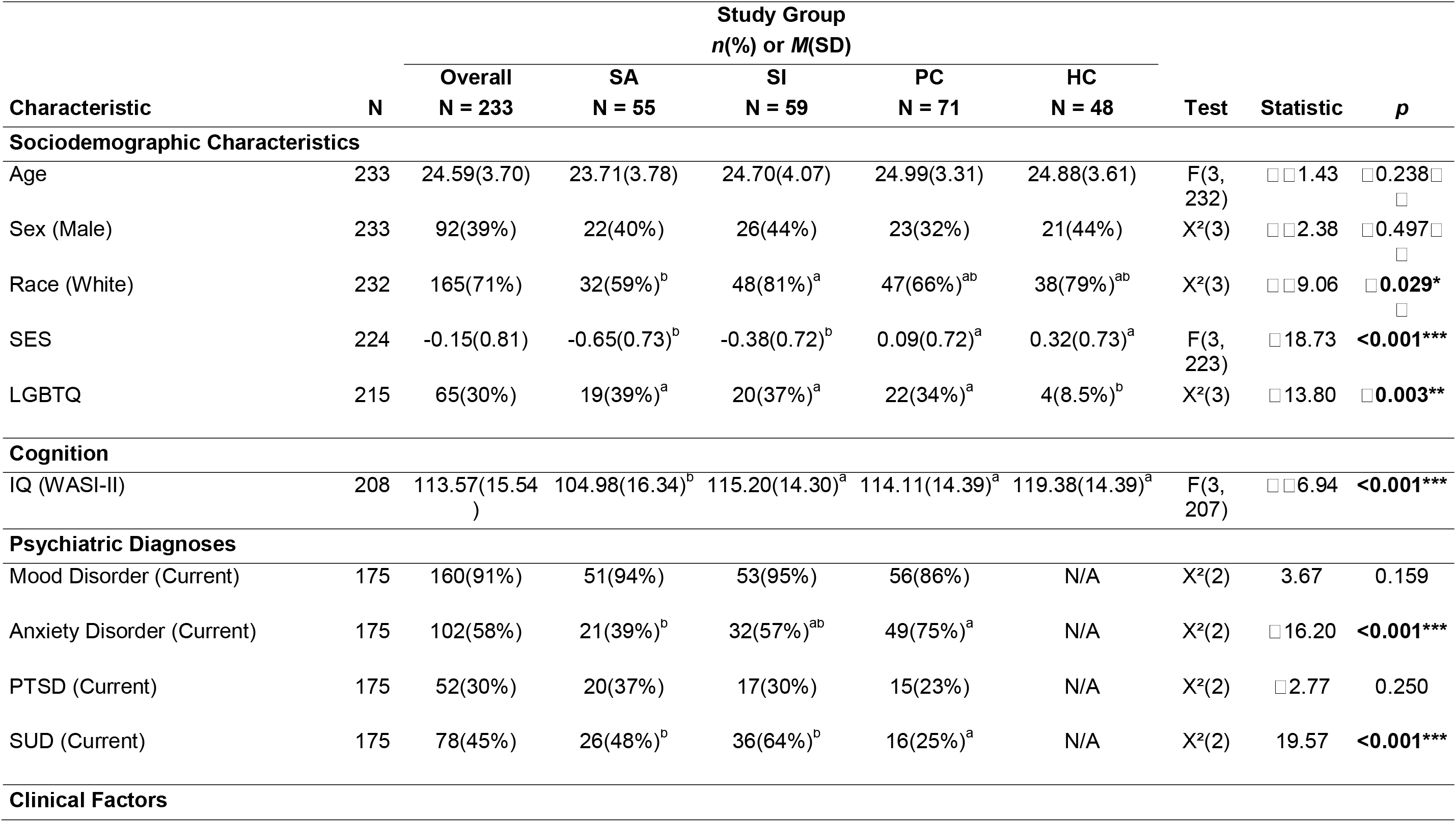
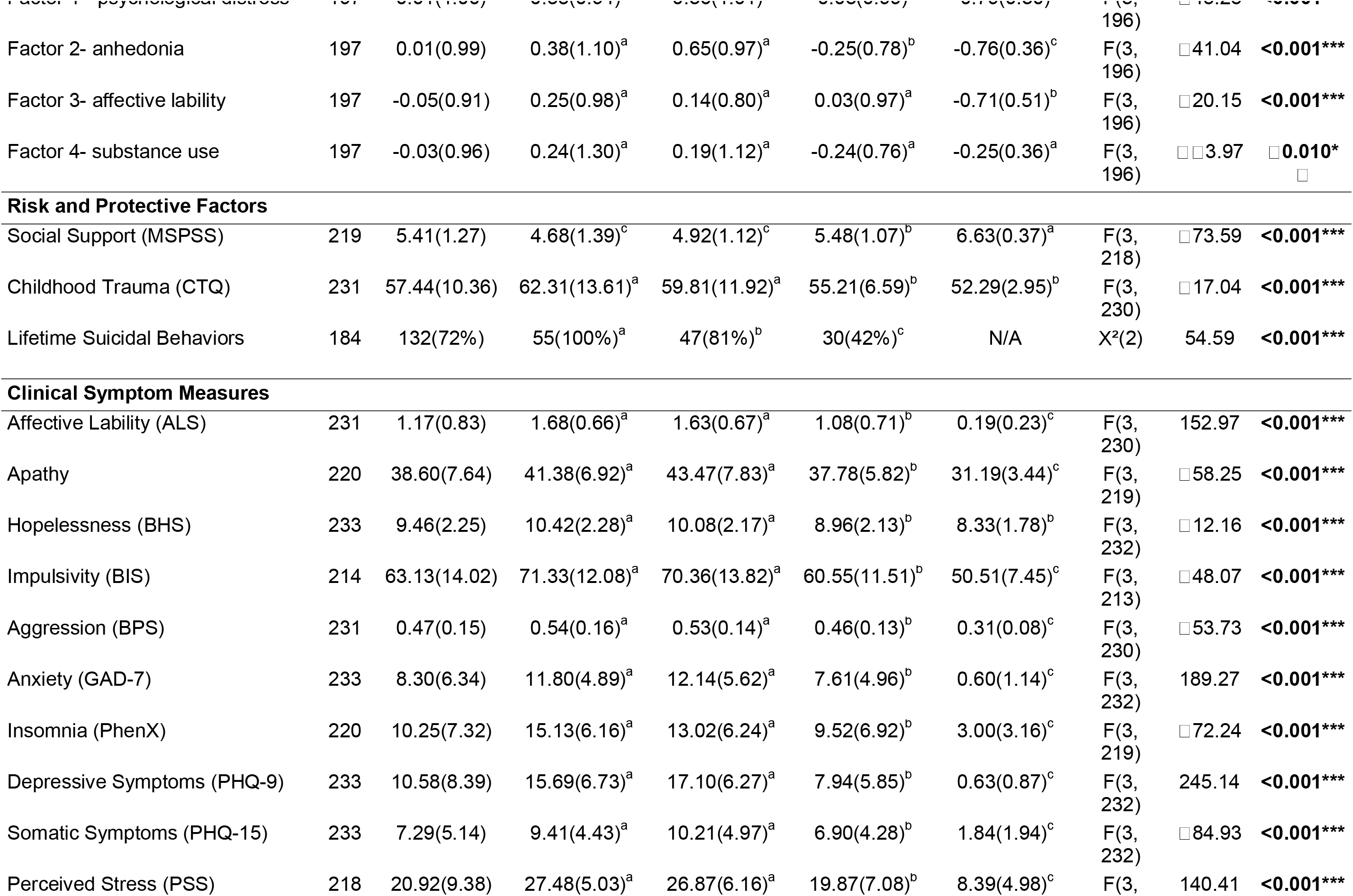
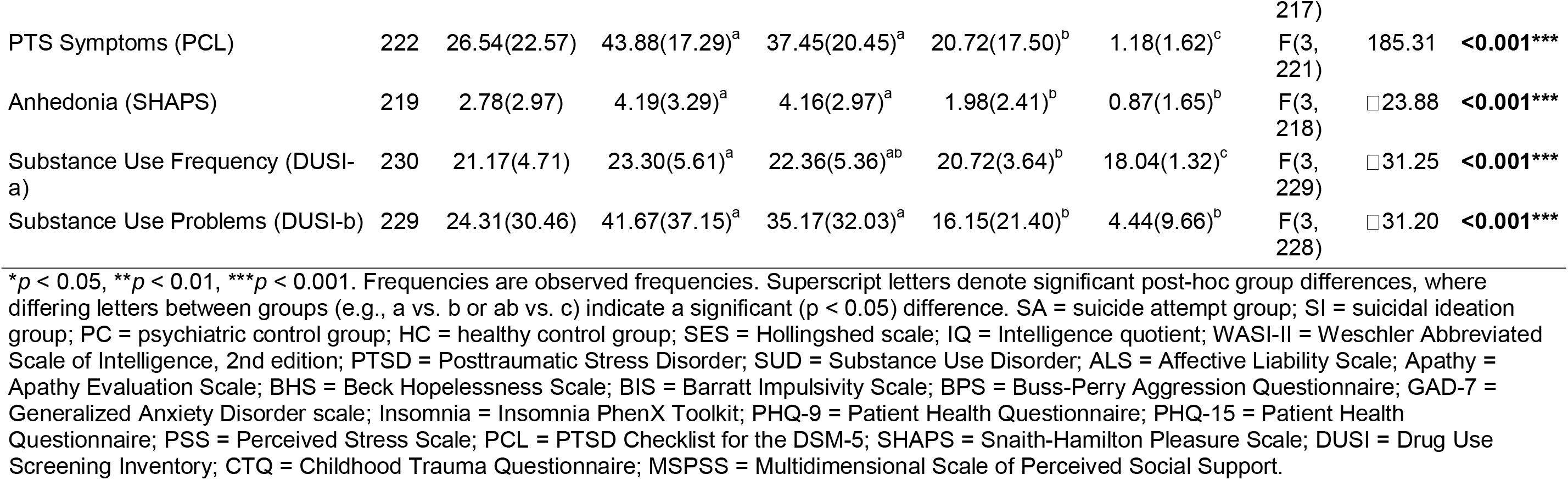
Sample Characteristics and Study Group Comparisons.

At baseline, the SA group had significantly lower IQ scores (M = 105.0, SD = 16.3) compared to all other groups. In addition, the SA group had higher rates of anxiety and substance use disorders compared to PC. In terms of lifetime history of suicidal behaviors, 100% (n = 55) of SA participants, 81% (n = 47) of SI, and 42% (n = 30) of PC reported such history.

In general, the SA and SI groups had similar levels of each of the clinical factors and higher levels of each clinical factor compared to the PC and HC groups. The same was true for the clinical symptom measures that were used in factor analyses. Among the risk and protective factors, the SA and SI had lower levels of perceived support and higher levels of childhood trauma compared to the PC and HC groups (**Table 1**).

### Group differences in Stressful Cups Task performance at baseline

#### Proportion of risky choices

There were no significant differences between groups in the proportion of risky choices under neutral or stress conditions or in the stress-minus-neutral difference after adjustment for multiple comparisons. Group differences also remained non-significant after controlling for clinical and sociodemographic covariates (**Supplementary Tables 5-6**).

#### EV sensitivity

There were no significant differences between groups in EV sensitivity under neutral or stress conditions or in the stress-minus-neutral difference after adjustment for multiple comparisons. Group differences remained non-significant after controlling for clinical and sociodemographic covariates (**Supplementary Tables 5-6**).

### Changes in Stressful Cups Task performance over time

#### Proportion of risky choices

For the group by time interactions, IPW analyses adjusting for attrition showed that SA participants had a significantly greater decline in the proportion of risky choices under stress over time relative to HC (**Supplementary Table 7)**, even when adjusting for covariates (β = −0.119, SE = 0.021, Tukey *p* < 0.001, Cohen’s *d* = 0.534; **Table 2**; **Figure 1)**. For the stress-minus-neutral difference, the SA by time interaction was also significant after controlling for covariates (β = −0.051, SE = 0.017, Tukey *p* = 0.019, Cohen’s *d* = 0.401; **Table 2**), indicating that SA participants tend to choose less riskier options under stress compared to neutral over time (See **Figure 2**). No other group by time interactions were significant after adjustment for multiple comparisons or controlling for covariates.

**Table 2.** Linear mixed effects models for differences between groups at changes in Cups task performance over time while adjusting for clinical and sociodemographic covariates IPW weighted analysis

| **Proportion of risky choices** |  |  |  |  |  |  |  |  |  |  |  |  |
|---|---|---|---|---|---|---|---|---|---|---|---|---|
| **Condition** | **Neutral (N = 202)** |  |  |  | **Stress (N = 192)** |  |  |  | **Difference (Stress − Neutral) (N = 189)** |  |  |  |
|  | β(SE) | *p* | Tukey p | Cohen’s d | β(SE) | *p* | Tukey p | Cohen’s d | β(SE) | *p* | Tukey p | Cohen’s d |
| PC vs HC | −0.009 (0.039) | 0.824 | 0.996 | 0.045 | −0.055 (0.046) | 0.228 | 0.621 | 0.249 | −0.019 (0.029) | 0.507 | 0.910 | 0.150 |
| SI vs HC | −0.023 (0.040) | 0.570 | 0.941 | 0.118 | −0.074 (0.047) | 0.112 | 0.383 | 0.335 | −0.083 (0.031) | 0.008 | 0.041 | 0.662 |
| SA vs HC | 0.010 (0.043) | 0.816 | 0.996 | 0.051 | 0.036 (0.049) | 0.471 | 0.888 | 0.161 | −0.007 (0.033) | 0.822 | 0.996 | 0.058 |
| Time | 0.015 (0.012) | 0.225 | — | 0.078 | 0.017 (0.013) | 0.182 | — | 0.078 | 0.005 (0.010) | 0.614 | — | 0.042 |
| PC × Time | 0.017 (0.020) | 0.393 | 0.826 | 0.088 | −0.005 (0.022) | 0.828 | 0.996 | 0.021 | −0.034 (0.017) | 0.050 | 0.200 | 0.267 |
| SI × Time | 0.022 (0.021) | 0.306 | 0.733 | 0.113 | 0.015 (0.021) | 0.476 | 0.890 | 0.068 | 0.005 (0.017) | 0.752 | 0.989 | 0.043 |
| SA × Time | −0.024 (0.017) | 0.168 | 0.510 | 0.121 | −0.119 (0.021) | **<0.001\*\*** | **<0.001\*** | 0.534 | −0.051 (0.017) | **0.004\*\*** | **0.019\*** | 0.401 |
| IQ | −0.004 (0.013) | 0.766 | — | 0.020 | −0.006 (0.016) | 0.707 | — | 0.027 | −0.010 (0.011) | 0.334 | — | 0.082 |
| Female | −0.071 (0.027) | 0.009 | — | 0.366 | — | — | — | — | — | — | — | — |
| Factor 2-Anhedonia | — | — | — | — | — | — | — | — | 0.025 (0.012) | 0.030 | — | 0.201 |
| **EV Sensitivity** |  |  |  |  |  |  |  |  |  |  |  |  |
| **Condition** | **Neutral (N = 202)** |  |  |  | **Stress (N = 192)** |  |  |  | **Difference (Stress − Neutral) (N = 189)** |  |  |  |
|  | β(SE) | *p* | Tukey p | Cohen’s d | β(SE) | *p* | Tukey p | Cohen’s d | β(SE) | *p* | Tukey p | Cohen’s d |
| PC vs HC | 0.019 (0.051) | 0.707 | 0.982 | 0.077 | 0.062 (0.057) | 0.277 | 0.694 | 0.230 | 0.020 (0.050) | 0.685 | 0.977 | 0.094 |
| SI vs HC | 0.059 (0.052) | 0.252 | 0.659 | 0.238 | 0.103 (0.057) | 0.072 | 0.270 | 0.380 | 0.015 (0.056) | 0.786 | 0.993 | 0.070 |
| SA vs HC | 0.048 (0.055) | 0.376 | 0.811 | 0.195 | 0.075 (0.061) | 0.215 | 0.599 | 0.276 | 0.078 (0.058) | 0.178 | 0.530 | 0.360 |
| Time | −0.007 (0.018) | 0.709 | — | 0.027 | 0.004 (0.020) | 0.850 | — | 0.014 | 0.011 (0.019) | 0.582 | — | 0.049 |
| PC × Time | 0.022 (0.029) | 0.438 | 0.864 | 0.089 | −0.010 (0.034) | 0.776 | 0.992 | 0.035 | −0.033 (0.033) | 0.326 | 0.760 | 0.150 |
| SI × Time | 0.031 (0.030) | 0.312 | 0.741 | 0.124 | 0.025 (0.033) | 0.454 | 0.875 | 0.091 | −0.003 (0.031) | 0.932 | 1.000 | 0.012 |
| SA × Time | 0.003 (0.025) | 0.887 | 0.999 | 0.014 | −0.026 (0.033) | 0.443 | 0.867 | 0.094 | −0.025 (0.032) | 0.429 | 0.858 | 0.116 |
| IQ | −0.126 (0.017) | <0.001 | — | 0.506 | −0.123 (0.019) | <0.001 | — | 0.453 | −0.003 (0.019) | 0.873 | — | 0.014 |
| Female | 0.096 (0.033) | 0.005 | — | 0.385 | 0.121 (0.038) | 0.002 | — | 0.443 | — | — | — | — |
| White | −0.135 (0.036) | **<0.001\*\*** | — | 0.545 | −0.103 (0.042) | 0.015 | — | 0.377 | — | — | — | — |
| SES | — | — | — | — | — | — | — | — | 0.074 (0.021) | <0.001 | — | 0.339 |
| Factor 2-Anhedonia | — | — | — | — | — | — | — | — | 0.046 (0.021) | 0.029 | — | 0.210 |

\*p < 0.05, \*\*p < 0.01, \*\*\*p < 0.001. SA = suicide attempt group; SI = suicidal ideation group; PC = psychiatric control group; HC = healthy control group; IQ = Intelligence Quotient, EV – Expected Value.

**Figure 1.**
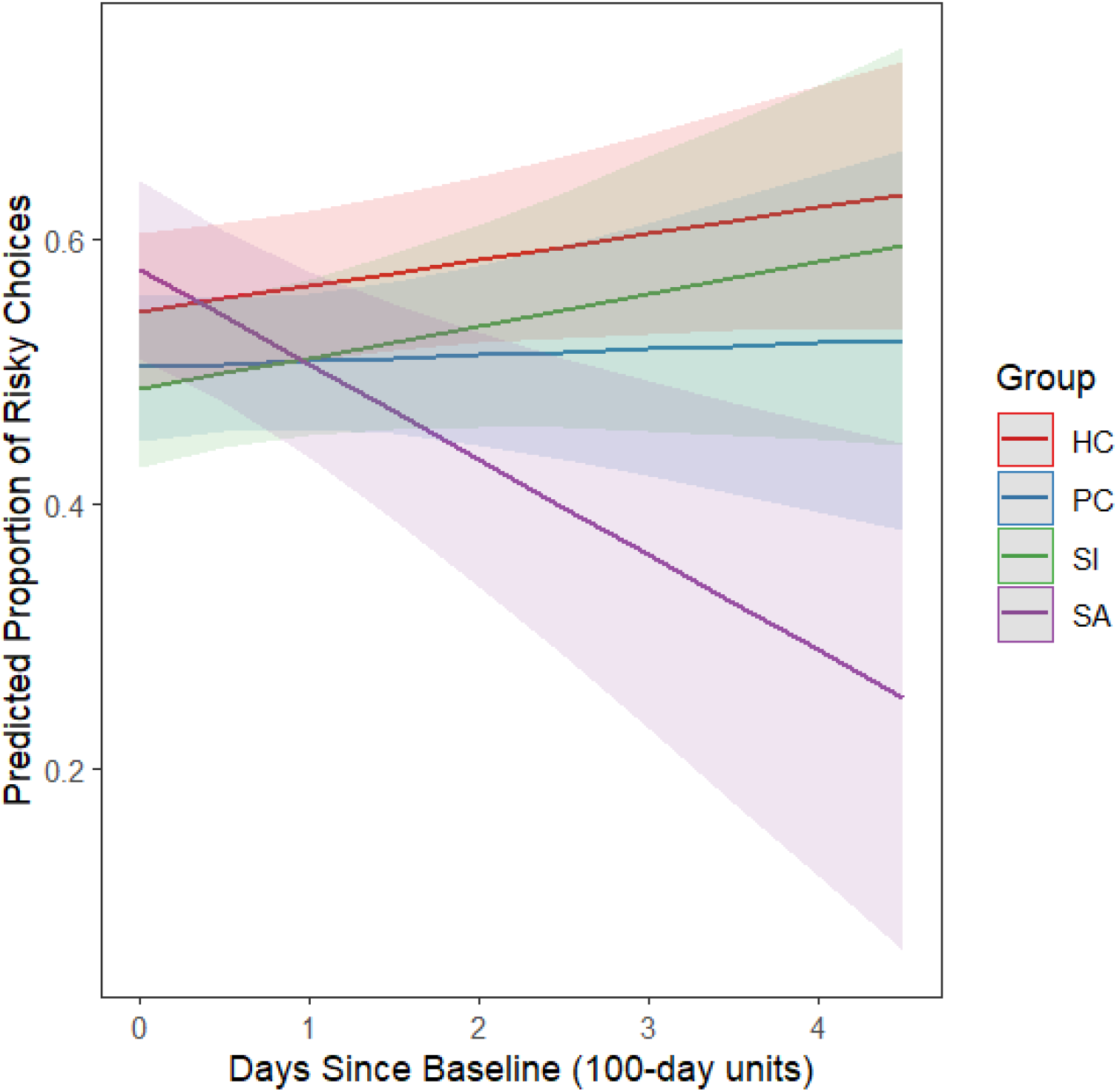
SA by time interaction on the changes in proportion of risky responses under stress over time

**Figure 2.**
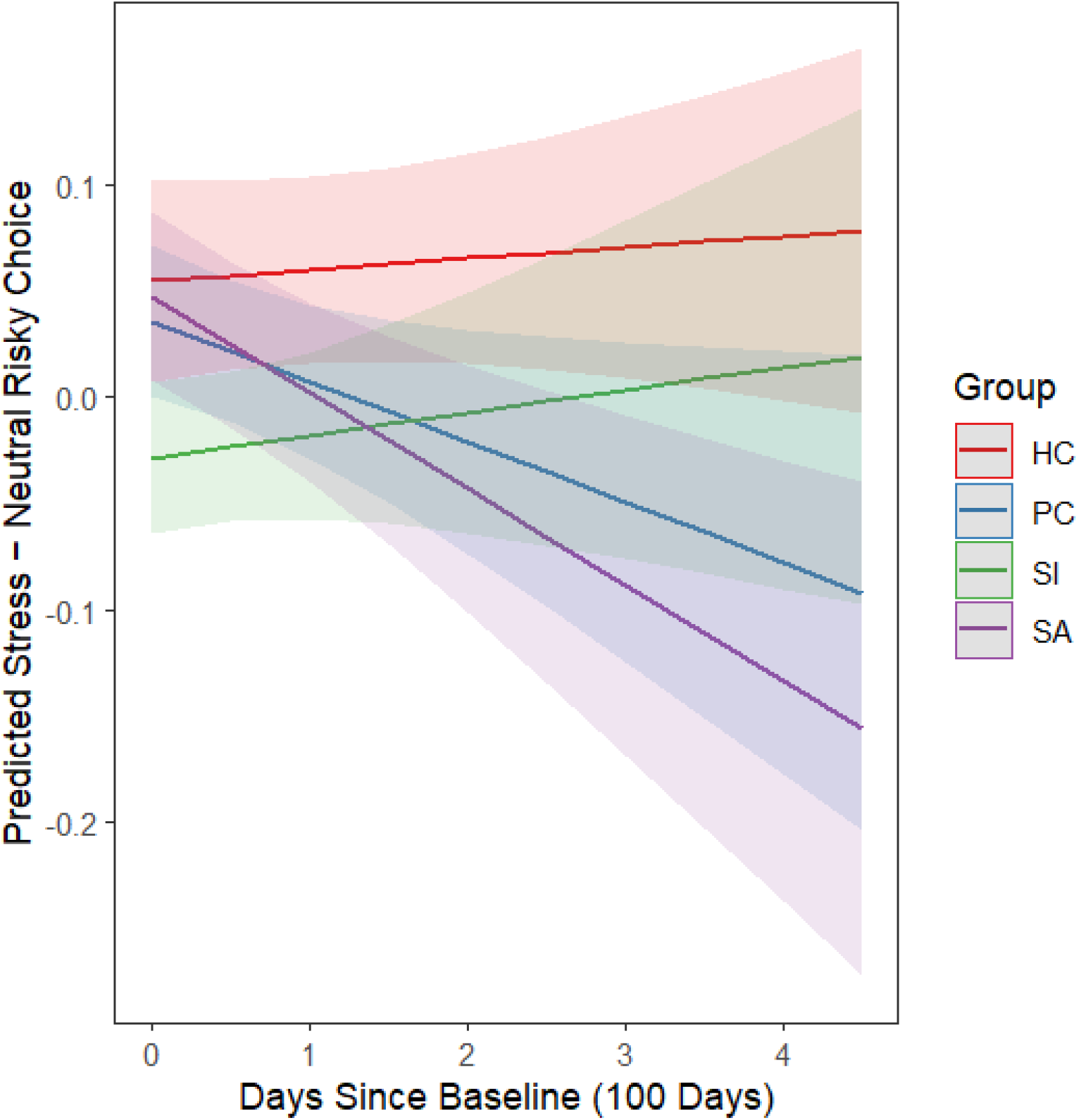
SA by time interaction on the changes in the difference between proportion of risky responses under stress vs. neutral over time

In the non-IPW sensitivity analyses, the SA by time interaction for risky choices under stress was replicated (β = −0.092, SE = 0.025, Tukey *p* = 0.002, Cohen’s *d* = 0.427; **Supplementary Table 8**), whereas the SA by time interaction for the stress-minus-neutral difference was not replicated (β = −0.024, SE = 0.019, Tukey *p* = 0.588, Cohen’s *d* = 0.202; **Supplementary Tables 8)**. An SA by time interaction on risky choices under neutral conditions was significant in the non-IPW sensitivity analysis (β = −0.056, SE = 0.020, Tukey *p* = 0.031, Cohen’s *d* = 0.294), although this interaction was not significant in the primary IPW analysis.

#### EV sensitivity

No significant interactions were observed for EV sensitivity under neutral or stress conditions or for the stress-minus-neutral difference after adjustment for multiple comparisons and controlling for covariates (**Supplementary Table 7)**.

### Association between biological measures and Stressful Cups Task performance at baseline

#### Proportion of risky choices

No associations between any biological measures (including HCC, CRP, IL6, IL10, IL-1ra and TNF-α) and the proportion of risky choices remained significant after correction for multiple comparisons and adjustment for clinical and sociodemographic covariates (**Supplementary Tables 9-10**).

#### EV sensitivity

No associations between any biological measures (including HCC, CRP, IL6, IL10, IL-1ra and TNF-α) and EV sensitivity remained significant after correction for multiple comparisons and adjustment for clinical and sociodemographic covariates (**Supplementary Tables 9-10**).

### Biological measures as predictors of changes in Stressful Cups task performance over time

#### Proportion of risky choices

For the main effects of the biological measures, higher CRP was associated with fewer risky choices under both neutral and stress conditions in the longitudinal IPW models (**Supplementary Table 11)**, and these main effects remained significant after controlling for clinical and sociodemographic covariates (neutral: β = −0.040, SE = 0.010, *p* < 0.001, Cohen’s *d* = 0.216; stress: β = −0.131, SE = 0.031, *p* < 0.001, Cohen’s *d* = 0.610; **Table 3**). Higher IL-6 also had a significant main effect on risky choices under the neutral condition, with higher IL-6 associated with fewer risky choices, which remained significant after adjustment for covariates (β = −0.080, SE = 0.018, *p* < 0.001, Cohen’s *d* = 0.442; **Table 3**).

**Table 3.** Linear mixed effects models for the association between changes in biomarkers and changes in Cups task performance over time while adjusting for clinical and sociodemographic covariates IPW weighted analysis

| Proportion of risky choices |  |  |  |  |  |  |  |
|---|---|---|---|---|---|---|---|
| **Condition** | **Neutral (N=177)** |  |  |  | **Neutral (N=165)** |  |  |
|  | β (SE) | p | Cohen's d |  | β (SE) | p | Cohen's d |
| CRP | −0.040 (0.010) | **<0.001**\*\*\* | 0.216 | IL-6 | −0.080 (0.018) | **<0.001**\*\*\* | 0.442 |
| Time | 0.014 (0.010) | 0.137 | 0.076 | Time | 0.009 (0.016) | 0.588 | 0.048 |
| CRP × Time | −0.000 (0.008) | 0.978 | 0.001 | IL-6 × Time | 0.006 (0.013) | 0.653 | 0.032 |
| PC vs HC | 0.004 (0.038) | 0.911 | 0.023 | PC vs HC | 0.021 (0.037) | 0.568 | 0.115 |
| SI vs HC | −0.023 (0.039) | 0.559 | 0.122 | SI vs HC | −0.028 (0.038) | 0.462 | 0.154 |
| SA vs HC | 0.018 (0.045) | 0.683 | 0.098 | SA vs HC | 0.018 (0.043) | 0.671 | 0.100 |
| IQ | 0.002 (0.014) | 0.908 | 0.009 | IQ | 0.005 (0.014) | 0.718 | 0.027 |
| BMI | 0.014 (0.015) | 0.379 | 0.073 | BMI | 0.025 (0.015) | 0.099 | 0.139 |
|  |  |  |  | Female | −0.078 (0.027) | **0.005**\*\* | 0.428 |
| **Condition** | **Stress (N=163)** |  |  |  | **Difference (Stress − Neutral) (N=150)** |  |  |
|  | β (SE) | p | Cohen's d |  | β (SE) | p | Cohen's d |
| CRP | −0.131 (0.031) | **<0.001**\*\*\* | 0.610 | IL-10 | 0.026 (0.026) | 0.329 | 0.205 |
| Time | 0.007 (0.012) | 0.547 | 0.034 | Time | 0.069 (0.022) | **0.002**\*\* | 0.554 |
| CRP × Time | −0.055 (0.028) | 0.051 | 0.254 | IL-10 × Time | −0.068 (0.018) | **<0.001**\*\*\* | 0.550 |
| PC vs HC | −0.043 (0.045) | 0.335 | 0.202 | PC vs HC | −0.035 (0.027) | 0.202 | 0.282 |
| SI vs HC | −0.064 (0.045) | 0.162 | 0.296 | SI vs HC | −0.086 (0.031) | **0.006**\*\* | 0.695 |
| SA vs HC | 0.026 (0.052) | 0.624 | 0.120 | SA vs HC | −0.055 (0.037) | 0.132 | 0.446 |
| IQ | −0.003 (0.017) | 0.852 | 0.015 | IQ | −0.010 (0.011) | 0.363 | 0.084 |
| BMI | 0.029 (0.019) | 0.119 | 0.137 | BMI | −0.003 (0.010) | 0.796 | 0.022 |
|  |  |  |  | Factor 2-Anhedonia | 0.036 (0.013) | **0.006**\*\* | 0.287 |

\*p < 0.05, \*\*p < 0.01, \*\*\*p < 0.001.

For the biomarker by time interactions, the IL-10 by time interaction for the stress-minus-neutral difference in proportion of risky choices remained significant after controlling for covariates (β = −0.068, SE = 0.018, *p* < 0.001, Cohen’s *d* = 0.550; **Table 3**). This result indicates that higher IL10 was associated with choosing lower risky options under stress compared to neutral over time (See **Figure 3**). No other biological measures (including HCC, IL-1ra and TNF-α) by time interactions remained significant after correction for multiple comparisons or adjustment for covariates (**Supplementary Table 11)**.

**Figure 3.**
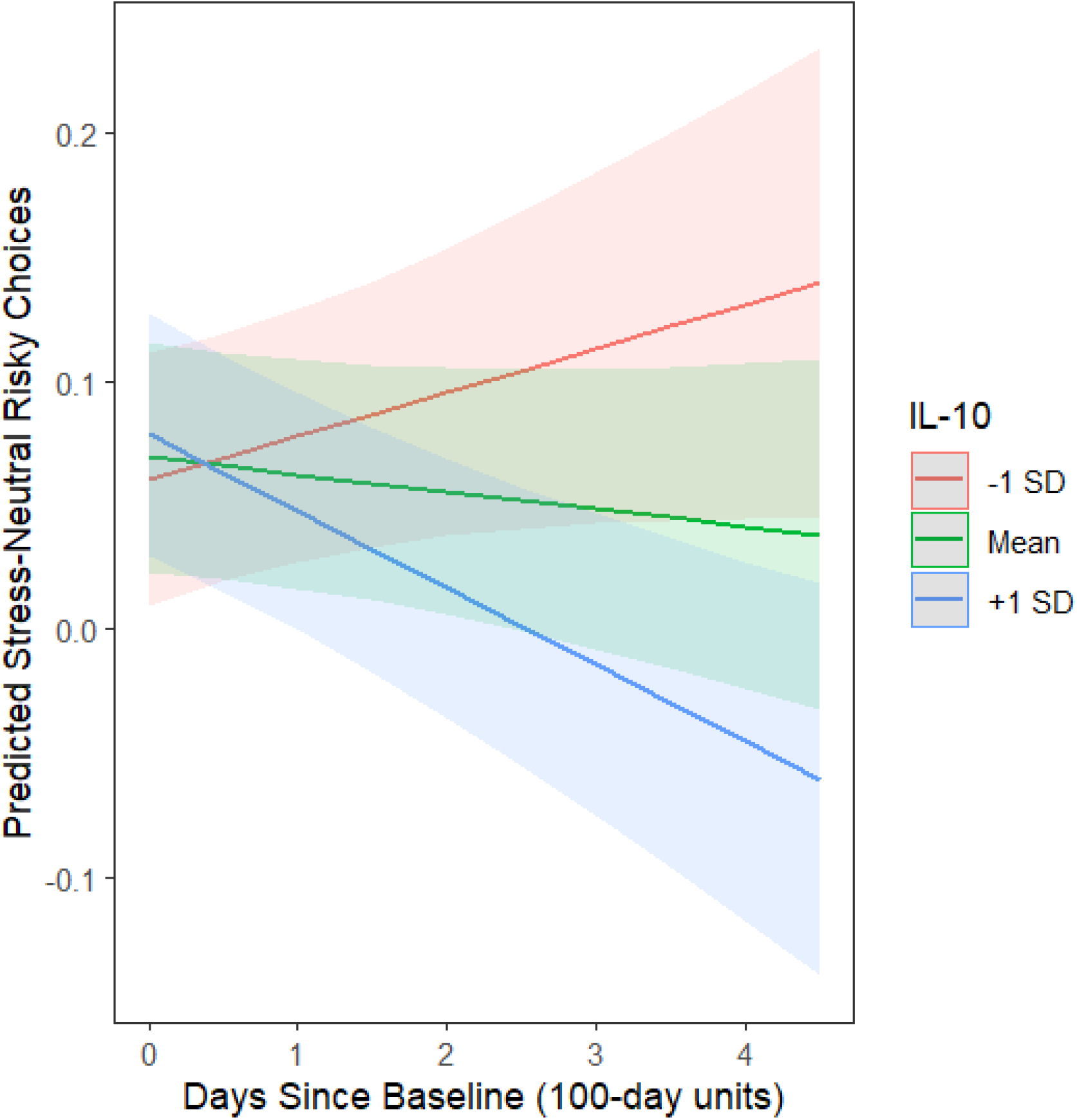
IL-10 by time interaction on changes in the difference between proportion of risky responses under stress vs. neutral over time

In the non-IPW sensitivity analyses, the main effects of CRP and IL-6 on risky choices under the neutral condition were replicated, whereas the IL-10 by Time interaction for the stress-minus-neutral difference was not replicated (**Supplementary Table 12)**.

#### EV sensitivity

Neither main effects of the biological measures (including HCC, CRP, IL6, IL10, IL-1ra and TNF-α) nor biomarker by time interactions for EV sensitivity remained significant after correction for multiple comparisons and adjustment for covariates (**Supplementary Table 11)**.

### Baseline Stressful Cups task performance predicting prospective Suicide Attempts and Suicidal Behaviors

#### Proportion of risky choices

The proportion of baseline risky choices under neutral or stress conditions or the stress-minus-neutral difference did not predict SA or SB outcomes after controlling for clinical covariates (**Table 4; Supplementary Table 13**).

**Table 4.**
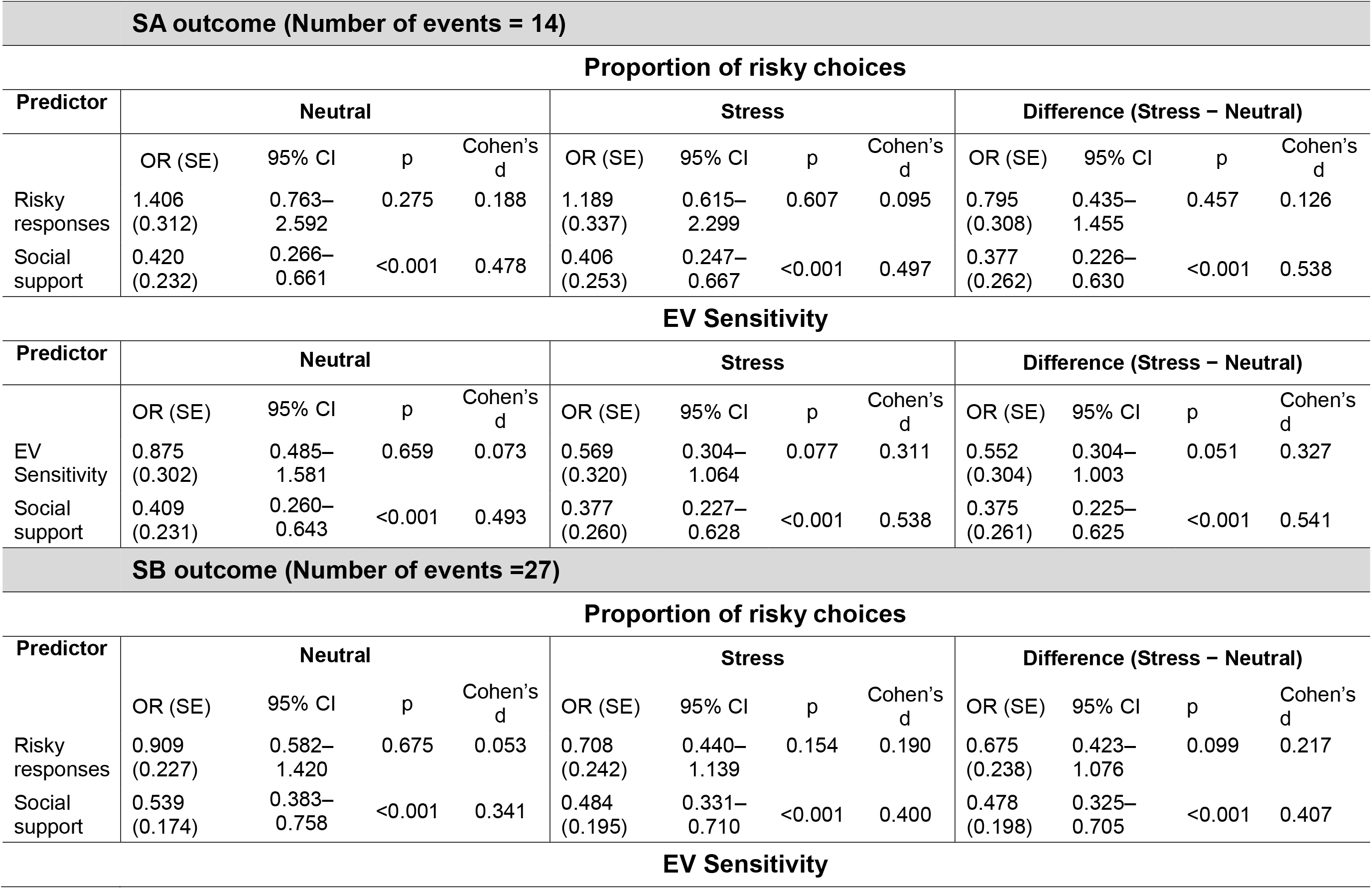
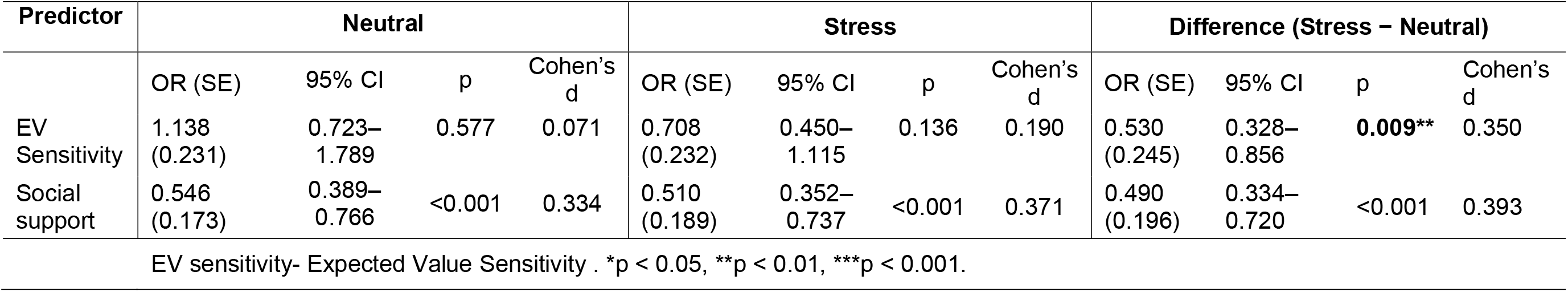
Logistic models for Cups performance predicting SA and SB outcomes adjusting for clinical covariates (social support)

|  |  |  |  |  |  |  |  |  |  |  |  |  |
|---|---|---|---|---|---|---|---|---|---|---|---|---|
|  | **SA outcome (Number of events = 14)** |  |  |  |  |  |  |  |  |  |  |  |
|  | **Proportion of risky choices** |  |  |  |  |  |  |  |  |  |  |  |
| **Predictor** | **Neutral** |  |  |  | **Stress** |  |  |  | **Difference (Stress − Neutral)** |  |  |  |
|  | OR (SE) | 95% CI | p | Cohen’s d | OR (SE) | 95% CI | p | Cohen’s d | OR (SE) | 95% CI | p | Cohen’s d |
| Risky responses | 1.406 (0.312) | 0.763–2.592 | 0.275 | 0.188 | 1.189 (0.337) | 0.615–2.299 | 0.607 | 0.095 | 0.795 (0.308) | 0.435–1.455 | 0.457 | 0.126 |
| Social support | 0.420 (0.232) | 0.266–0.661 | <0.001 | 0.478 | 0.406 (0.253) | 0.247–0.667 | <0.001 | 0.497 | 0.377 (0.262) | 0.226–0.630 | <0.001 | 0.538 |
|  | **EV Sensitivity** |  |  |  |  |  |  |  |  |  |  |  |
| **Predictor** | **Neutral** |  |  |  | **Stress** |  |  |  | **Difference (Stress − Neutral)** |  |  |  |
|  | OR (SE) | 95% CI | p | Cohen’s d | OR (SE) | 95% CI | p | Cohen’s d | OR (SE) | 95% CI | p | Cohen’s d |
| EV Sensitivity | 0.875 (0.302) | 0.485–1.581 | 0.659 | 0.073 | 0.569 (0.320) | 0.304–1.064 | 0.077 | 0.311 | 0.552 (0.304) | 0.304–1.003 | 0.051 | 0.327 |
| Social support | 0.409 (0.231) | 0.260–0.643 | <0.001 | 0.493 | 0.377 (0.260) | 0.227–0.628 | <0.001 | 0.538 | 0.375 (0.261) | 0.225–0.625 | <0.001 | 0.541 |
|  | **SB outcome (Number of events =27)** |  |  |  |  |  |  |  |  |  |  |  |
|  | **Proportion of risky choices** |  |  |  |  |  |  |  |  |  |  |  |
| **Predictor** | **Neutral** |  |  |  | **Stress** |  |  |  | **Difference (Stress − Neutral)** |  |  |  |
|  | OR (SE) | 95% CI | p | Cohen’s d | OR (SE) | 95% CI | p | Cohen’s d | OR (SE) | 95% CI | p | Cohen’s d |
| Risky responses | 0.909 (0.227) | 0.582–1.420 | 0.675 | 0.053 | 0.708 (0.242) | 0.440–1.139 | 0.154 | 0.190 | 0.675 (0.238) | 0.423–1.076 | 0.099 | 0.217 |
| Social support | 0.539 (0.174) | 0.383–0.758 | <0.001 | 0.341 | 0.484 (0.195) | 0.331–0.710 | <0.001 | 0.400 | 0.478 (0.198) | 0.325–0.705 | <0.001 | 0.407 |
|  | **EV Sensitivity** |  |  |  |  |  |  |  |  |  |  |  |
| **Predictor** | **Neutral** |  |  |  | **Stress** |  |  |  | **Difference (Stress − Neutral)** |  |  |  |
|  | OR (SE) | 95% CI | p | Cohen’s d | OR (SE) | 95% CI | p | Cohen’s d | OR (SE) | 95% CI | p | Cohen’s d |
| EV Sensitivity | 1.138 (0.231) | 0.723–1.789 | 0.577 | 0.071 | 0.708 (0.232) | 0.450–1.115 | 0.136 | 0.190 | 0.530 (0.245) | 0.328–0.856 | **0.009**\*\* | 0.350 |
| Social support | 0.546 (0.173) | 0.389–0.766 | <0.001 | 0.334 | 0.510 (0.189) | 0.352–0.737 | <0.001 | 0.371 | 0.490 (0.196) | 0.334–0.720 | <0.001 | 0.393 |

EV sensitivity- Expected Value Sensitivity . \*p < 0.05, \*\*p < 0.01, \*\*\*p < 0.001.

#### EV sensitivity

Lower expected value sensitivity under stress relative to neutral conditions was associated with higher odds of subsequent suicidal behavior (OR = 0.530, SE= 0.245, 95% CI [0.328, 0.856], p = 0.009) while adjusting for clinical covariates. (OR = 0.553, SE=0.223, 95% CI [0.358, 0.856], *p* = 0.008; **Supplementary Table 13**) and remained significant after controlling for social support (OR = 0.530, SE= 0.245, 95% CI [0.328, 0.856], *p* = 0.009; **Table 4**). Thus, individuals who showed a decline in their ability to distinguish between advantageous and disadvantageous risky choices under stress were more likely to engage in SB during follow-up. None of the other outcomes for EV sensitivity at baseline remained significant predictors of SA or SB after controlling for clinical covariates (**Table 4; Supplementary Table 13**).

### Time-to-Onset of Suicide Attempts and Suicidal Behaviors

#### Proportion of risky choices

No associations between proportion of risky choices under neutral or stress conditions or the stress-minus-neutral difference significantly predicted time to onset of suicidal attempt or behavior after controlling for clinical covariates (**Supplementary Tables 14-15)**.

#### EV sensitivity

The stress-minus-neutral difference in proportion of risky choices significantly predicted time to onset of SA in the unadjusted model (HR = 0.619, 95% CI [0.401, 0.956], p=0.031). However, this result did not remain significant after controlling for clinical covariates (HR = 0.625, 95% CI [0.367, 1.063], p=0.083) (**Supplementary Tables 14-15**).

## Discussion

Our findings highlight the importance of considering stress when examining decision-making processes and risk for suicidal behavior among young adults. Participants with a history of suicide attempts showed greater reductions in risky choices under stress over time compared to healthy controls, suggesting distinct longitudinal patterns of decision-making among individuals with a history of suicide attempts. Moreover, greater reductions in expected value sensitivity under stress relative to neutral conditions prospectively predicted subsequent suicidal behavior. Together, these findings suggest that reduced risk-taking and difficulty distinguishing between advantageous and disadvantageous choices under stress may be relevant to subsequent suicide risk. Biological stress responses were also associated with decision-making. Higher levels of inflammation, particularly CRP, IL-6 and IL-10, were associated with greater reductions in risky choices over time under stress.

These findings should be interpreted in the context of several limitations. First, the high attrition rate and clinical differences between participants retained and those lost to follow-up may influence the findings. However, retention challenges are common in similarly high-risk clinical populations ^51^. To address potential attrition-related bias, all longitudinal analyses used inverse probability weighting (IPW). Second, participants who did not complete the Stressful Cups task reported higher levels of clinical symptoms compared to those who did. As these factors may influence both decision-making and biological markers, this may have biased our results towards the null hypotheses, reducing our ability to detect significant association between risky decision-making and suicide-related outcomes. However, we also controlled for these variables in our analyses. Third, the sample size limited our ability to examine whether longitudinal associations between biological markers and risky decision-making differed among participants with a history of suicide attempts, those with suicidal ideation, psychiatric controls, and healthy controls using three-way interactions between biomarkers, time, and group. Fourth, the racial composition of the sample, which was predominantly White (72.3%), may limit the extent to which these findings generalize to more diverse populations.

Contrary to our hypothesis, our results indicate that longitudinally, participants with a history of suicide attempt showed a greater decline in risky choices under stress over time relative to healthy controls. Moreover, participants with a history of suicide attempt became increasingly less likely to choose risky options under stress relative to neutral conditions over time compared with healthy controls. These findings are consistent with prior evidence of greater avoidance of risk and loss among individuals with a history of suicide attempt. One study found that depressed suicide attempters demonstrated greater risk aversion when facing potential losses and greater loss aversion than non-suicidal depressed patients and healthy controls ^7^. Similarly, another study found greater loss aversion among depressed suicide attempters than both non-suicidal depressed patients and healthy controls, as well as greater psychological pain avoidance among attempters ^52^. Furthermore, another study found that adolescents with a history of suicide attempts selected lower risky options on the Iowa Gambling Task than depressed adolescents without a history of suicide attempts and healthy controls ^53^. In contrast, a substantial literature has found greater disadvantageous or risky decision-making among suicide attempters. Meta-analyses have shown that SA make riskier choices and perform more poorly on the IGT than both psychiatric and healthy controls, and this pattern has also been demonstrated among adolescent suicide attempters ^9^. Importantly, a longitudinal study demonstrated that lower loss aversion predicted suicide attempts over four months among adolescents ^54^. One possible explanation for these inconsistent findings is the role of stress on decision making. Individuals with a history of suicide attempts may not necessarily demonstrate a generalized tendency toward either greater risk-taking or risk avoidance, rather, stress may shift decision-making toward avoidance of uncertain or potentially aversive outcomes. In the context of theories conceptualizing suicidal behavior as an escape from intolerable distress ^55^, stress may strengthen the tendency to select an immediately safer and more certain option. To our knowledge, this is the first longitudinal study to show that participants with a history of suicide attempt demonstrate increasing avoidance of risky choices specifically under stress.

Our findings indicate that greater decreases in EV sensitivity under stress relative to neutral conditions were associated with higher odds of subsequent suicidal behavior, even after adjustment for clinical covariates. This finding is consistent with prior research suggesting that difficulty distinguishing between options with different expected values may be associated with suicidal behavior. For example, a prior study indicated that suicide attempters, compared to those with ideation, non-suicidal depressed patients, and healthy controls, displayed abnormally long decision times when choosing between similarly valued options and were less able to distinguish between the best and second-best options ^12^. A later study suggested that suicidal behavior may arise, in part, when individuals experiencing a crisis have difficulty learning and representing the values of alternative actions and making advantageous choices based on those values ^56^.

For biological stress responses, our findings suggest that higher levels of inflammatory markers (CRP and IL6) are associated with reduced risky decision-making cross-sectionally, particularly under stress. Longitudinally, higher IL-10 was associated with greater reductions in risky choices under stress relative to neutral conditions over time. These findings are consistent with a previous study demonstrating that stress-induced increases in IL-6 were associated with reduced risk tolerance in pursuit of rewards among adolescents with greater early-life adversity ^57^. However, they contrast with studies linking higher inflammatory markers, including CRP and IL-6, to more impulsive decision-making in non-clinical samples ^5^. Although we did not identify previous studies directly examining IL-10 in relation to risky decision-making, evidence regarding its association with cognitive functioning has been mixed. Higher IL-10 has been associated with better cognitive and psychomotor performance in patients with major depression^58^, whereas another study of patients with depression found no significant association ^59^.

Although HCC was not significantly associated with risky decision-making under either neutral or stress conditions, this finding is consistent with previous research reporting no significant association between HCC and risky decision-making in healthy adults ^60^. However, other studies have linked lower HCC to greater risky decision-making among healthy adolescents ^19^, while experimental studies have demonstrated that cortisol administration can increase risk-taking ^4,16^. Together, these mixed findings suggest that the relationship between cortisol and risky decision-making remains complex and warrants further investigation, particularly among individuals at elevated risk for suicidal behavior.

In summary, this study is the first to prospectively examine risky decision-making and EV sensitivity under stress in young adults at risk for suicidal behavior. We found that participants with a history of suicide attempt showed greater reductions in risky choices under stress over time, highlighting the importance of considering stress when examining longitudinal changes in decision-making. Moreover, greater reductions in EV sensitivity under stress relative to neutral conditions prospectively predicted subsequent suicidal behaviors. This suggests that difficulty adjusting choices according to expected value under stress may be relevant to future suicide risk. Inflammatory markers (IL-10) were also associated with decision-making, with higher levels associated with decreases in risky choices over time, particularly under stress, highlighting the potential role of inflammation in shaping stress-related decision-making. Clinically, these findings suggest that stress may strengthen avoidance responses aimed at escaping distress while also impairing the ability to distinguish advantageous from disadvantageous risky choices. Consistent with theories conceptualizing suicidal behavior as an escape from intolerable distress ^55^, these changes may help explain how decision-making under stress relates to subsequent suicidal behavior. Together, these findings highlight the importance of assessing decision-making under stress and suggest that examining inflammatory and biological stress responses alongside decision-making may help identify mechanisms underlying vulnerability to suicidal behavior.

## Data Availability

All data produced in the present study are available upon reasonable request to the authors

