## Supplemental tables 1-15 for "Decision-Making Under Stress, Biological Stress Responses, and Risk for Suicidal Behavior in Young Adults"

**Supplementary Tables**

**Supplementary Table 1. Group comparisons between participants who did vs. did not complete the Stressful Cups task at baseline**

| **Characteristic** | **Did not complete the Stressful Cups task** | **Completed the Stressful Cups task** | **Test** | **Statistic** | **p** |
| --- | --- | --- | --- | --- | --- |
| N | 164 | 233 |  |  |  |
| Group, n (%) |  |  | χ²(3) | 10.670 | **0.014*** |
| HC | 15 (9.1)^a^ | 48 (20.6)^b^ |  |  |  |
| PC | 52 (34.9)^a^ | 71 (38.4)^a^ |  |  |  |
| SI | 56 (37.6)^a^ | 59 (31.9)^a^ |  |  |  |
| SA | 41 (27.5)^a^ | 55 (29.7)^a^ |  |  |  |
| **Sociodemographic characteristics** | | | | | |
| Age, M (SD) | 24.07 (3.62) | 24.59 (3.70) | t (395) | -1.395 | 0.164 |
| Sex (male), n (%) | 63 (38.4) | 92 (39.5) | Χ²(1) | 0.046 | 0.912 |
| Race (white), n (%) | 122 (74.8) | 165 (71.1) | Χ²(1) | 0.669 | 0.482 |
| SES, M (SD) | -0.20 (0.77) | -0.15 (0.81) | t (347) | -0.478 | 0.633 |
| LGBTQ, n (%) | 53 (44.9) | 65 (30.2) | Χ²(1) | 7.179 | **0.010*** |
| **Cognition** | | | | | |
| FSIQ / IQ, M (SD) | 109.75 (17.14) | 113.57 (15.54) | t (265) | -1.631 | 0.104 |
| **Psychiatric Diagnosis** | | | | | |
| Mood disorder (current), n (%) | 117 (79.1) | 160 (72.7) | Χ²(1) | 1.903 | 0.209 |
| Anxiety disorder (current), n (%) | 60 (40.5) | 102 (46.4) | Χ²(1) | 1.217 | 0.319 |
| PTSD (current), n (%) | 28 (18.9) | 52 (23.6) | Χ²(1) | 1.157 | 0.344 |
| AUD/SUD (current), n (%) | 80 (54.1) | 105 (47.7) | Χ²(1) | 1.417 | 0.278 |
| **Clinical Factors** | | | | | |
| **Risk and Protective Factors** | | | | | |
| Social support (MSPSS), M (SD) | 5.29 (1.25) | 5.41 (1.27) | t (346) | -0.842 | 0.400 |
| Childhood trauma (CTQ), M (SD) | 60.07 (11.73) | 57.44 (10.36) | t (378) | 2.296 | **0.022*** |
| Lifetime SB, n (%) | 108 (67.1) | 132 (56.9) | Χ²(1) | 3.729 | 0.053 |
| **Clinical Symptom Measures** | | | | | |
| Affective lability (ALS), M (SD) | 1.45 (0.84) | 1.17 (0.83) | t (378) | 3.143 | **0.002*** |
| Apathy, M (SD) | 41.90 (8.39) | 38.60 (7.64) | t (348) | 3.772 | **<0.001*** |
| Hopelessness (BHS), M (SD) | 10.27 (2.36) | 9.46 (2.25) | t (384) | 3.403 | **0.001*** |
| Impulsivity (BIS), M (SD) | 67.61 (14.17) | 63.13 (14.02) | t (343) | 2.866 | **0.004*** |
| Aggression (BPS), M (SD) | 0.49 (0.17) | 0.47 (0.15) | t (379) | 1.529 | 0.127 |
| Anxiety (GAD-7), M (SD) | 10.70 (6.44) | 8.30 (6.34) | t (386) | 3.635 | **<0.001*** |
| Insomnia, M (SD) | 12.43 (7.53) | 10.25 (7.32) | t (347) | 2.649 | **0.008*** |
| Depressive symptoms (PHQ-9), M (SD) | 12.88 (8.12) | 10.58 (8.39) | t (386) | 2.675 | **0.008*** |
| Somatic symptoms (PHQ-15), M (SD) | 8.63 (5.50) | 7.29 (5.14) | t (386) | 2.448 | **0.015*** |
| Perceived stress (PSS), M (SD) | 23.21 (7.71) | 20.92 (9.38) | t (341) | 2.308 | **0.022*** |
| PTS symptoms (PCL), M (SD) | 34.67 (21.97) | 26.54 (22.57) | t (365) | 3.407 | **0.001*** |
| Anhedonia (SHAPS), M (SD) | 3.26 (3.19) | 2.78 (2.97) | t (342) | 1.379 | 0.169 |
| Substance use frequency (DUSI-a), M (SD) | 22.04 (5.44) | 21.17 (4.71) | t (373) | 1.642 | 0.101 |
| Substance use problems (DUSI-b), M (SD) | 28.65 (30.89) | 24.31 (30.46) | t (372) | 1.337 | 0.182 |

**Supplementary Table 2. Comparisons between those who were retained vs. not retained from the study**

| **Characteristic** | **Non-retained** | **Retained** | **Test** | **Statistic** | **P** |
| --- | --- | --- | --- | --- | --- |
| N | 99 | 134 |  |  |  |
| Group, n (%) |  |  | χ²(3) | 23.429 | **<0.001***** |
| HC | 11 (11.1)^a^ | 37 (27.6)^b^ |  |  |  |
| PC | 23 (23.2)^a^ | 48 (35.8) ^b^ |  |  |  |
| SI | 29 (29.3))^a^ | 30 (22.4))^a^ |  |  |  |
| SA | 36 (36.4))^a^ | 19 (14.2) ^b^ |  |  |  |
| **Sociodemographic characteristics** | | | | | |
| Age, M (SD) | 24.22 (3.69) | 24.87 (3.69) | t (231) | -1.319 | 0.189 |
| Sex (male), n (%) | 49 (49.5) | 43 (32.1) | Χ²(1) | 7.219 | **0.011*** |
| Race (white), n (%) | 67 (68.4) | 98 (73.1) | Χ²(1) | 0.626 | 0.519 |
| SES, M (SD) | -0.50 (0.70) | 0.08 (0.79) | t (222) | -5.613 | **<0.001***** |
| LGBTQ, n (%) | 25 (27.8) | 40 (32.0) | Χ²(1) | 0.442 | 0.607 |
| **Cognition** | | | | | |
| FSIQ / IQ, M (SD) | 105.07 (13.92) | 119.10 (14.00) | t (206) | -7.079 | **<0.001***** |
| **Psychiatric Diagnosis** | | | | | |
| Mood disorder (current), n (%) | 77 (84.6) | 83 (64.3) | Χ²(1) | 11.058 | **0.002**** |
| Anxiety disorder (current), n (%) | 35 (38.5) | 67 (51.9) | Χ²(1) | 3.897 | 0.066 |
| PTSD (current), n (%) | 16 (17.6) | 36 (27.9) | Χ²(1) | 3.151 | 0.107 |
| AUD/SUD (current), n (%) | 56 (61.5) | 49 (38.0) | Χ²(1) | 11.866 | **0.001**** |
| **Risk and Protective Factors** | | | | | |
| Social support (MSPSS), M (SD) | 5.24 (1.14) | 5.52 (1.35) | t(217) | -1.592 | 0.113 |
| Childhood trauma (CTQ), M (SD) | 58.38 (11.27) | 56.76 (9.64) | t(229) | 1.175 | 0.241 |
| Lifetime SB, n (%) | 68 (68.7) | 64 (48.1) | Χ²(1) | 9.789 | **0.003**** |
| **Clinical Symptom Measures** | | | | | |
| Affective lability (ALS), M (SD) | 1.42 (0.76) | 0.99 (0.84) | t (229) | 4.065 | **<0.001**** |
| Apathy, M (SD) | 39.88 (7.75) | 37.73 (7.46) | t (218) | 2.062 | **0.040*** |
| Hopelessness (BHS), M (SD) | 10.20 (2.38) | 8.91 (1.98) | t(231) | 4.513 | **<0.001***** |
| Impulsivity (BIS), M (SD) | 67.58 (12.99) | 60.32 (13.97) | t(212) | 3.807 | **<0.001***** |
| Aggression (BPS), M (SD) | 0.50 (0.15) | 0.44 (0.15) | t(229) | 3.164 | **0.002**** |
| Anxiety (GAD-7), M (SD) | 10.24 (5.89) | 6.87 (6.29) | t(231) | 4.159 | **<0.001***** |
| Insomnia, M (SD) | 12.11 (6.82) | 8.99 (7.40) | t(218) | 3.160 | **0.002**** |
| Depressive symptoms (PHQ-9), M (SD) | 13.18 (8.17) | 8.67 (8.07) | t(231) | 4.200 | **<0.001***** |
| Somatic symptoms (PHQ-15), M (SD) | 8.25 (4.66) | 6.58 (5.38) | t(231) | 2.484 | **0.014*** |
| Perceived stress (PSS), M (SD) | 23.86 (8.98) | 18.90 (9.14) | t(216) | 3.961 | **<0.001***** |
| PTS symptoms (PCL), M (SD) | 33.13 (21.22) | 21.61 (22.37) | t(220) | 3.878 | **<0.001***** |
| Anhedonia (SHAPS), M (SD) | 3.14 (3.00) | 2.54 (2.94) | t(217) | 1.480 | 0.140 |
| Substance use frequency (DUSI-a), M (SD) | 22.15 (5.44) | 20.44 (3.95) | t(228) | 2.766 | **0.006**** |
| Substance use problems (DUSI-b), M (SD) | 33.67 (33.00) | 17.30 (26.43) | t(227) | 4.167 | **<0.001***** |

**Supplementary Table 3. Factor Loadings from Exploratory Factor Analysis at Baseline**

| **Clinical Symptom Measure** | **Factor 1**  **Psychological Distress** | **Factor 2 Anhedonia** | **Factor 3**  **Affective Lability** | **Factor 4 Substance Use** |
| --- | --- | --- | --- | --- |
| Affective Lability (ALS) | 0.568 | 0.295 | **0.591** | 0.239 |
| Apathy | 0.302 | **0.734** | 0.122 | -0.005 |
| Hopelessness (BHS) | 0.245 | 0.194 | **0.378** | 0.207 |
| Impulsivity (BIS) | 0.369 | **0.429** | 0.375 | 0.374 |
| Aggression (BPS) | 0.298 | 0.167 | **0.724** | 0.275 |
| Anxiety (GAD-7) | **0.710** | 0.462 | 0.283 | 0.142 |
| Insomnia (PhenX) | **0.561** | 0.343 | 0.342 | 0.218 |
| Depressive Symptoms (PHQ-9) | **0.662** | 0.627 | 0.235 | 0.169 |
| Somatic Symptoms (PHQ-15) | **0.748** | 0.279 | 0.210 | 0.156 |
| Perceived Stress (PSS) | 0.477 | **0.564** | 0.492 | 0.128 |
| PTS Symptoms (PCL) | **0.540** | 0.393 | 0.374 | 0.279 |
| Anhedonia (SHAPS) | 0.136 | **0.631** | 0.084 | 0.129 |
| Substance Use Frequency (DUSI-a) | 0.305 | 0.060 | 0.177 | **0.586** |
| Substance Use Problems (DUSI-b) | 0.108 | 0.185 | 0.224 | **0.948** |
| For each measure, the strongest factor loading is in bold. Eigenvalues: Factor 1 = 3.45, Factor 2 = 2.04, Factor 3 = 1.99, Factor 4 = 1.74. ALS = Affective Liability Scale; Apathy = Apathy Evaluation Scale; BHS = Beck Hopelessness Scale; BIS = Barratt Impulsivity Scale; BPS = Buss-Perry Aggression Questionnaire; GAD-7 = Generalized Anxiety Disorder scale; Insomnia = Insomnia PhenX Toolkit; PHQ-9 = Patient Health Questionnaire; PHQ-15 = Patient Health Questionnaire; PSS = Perceived Stress Scale; PCL = PTSD Checklist for the DSM-5; SHAPS = Snaith-Hamilton Pleasure Scale; DUSI = Drug Use Screening Inventory. *Note*. Bartlett’s test of sphericity was significant, χ²(120) = 2,639.03, p < .001, and the Kaiser–Meyer–Olkin (KMO) measure of sampling adequacy was 0.93, supporting the suitability of the data for factor analysis. | | | | |

**Supplementary Table 4. Measurement Invariance of Confirmatory Factor Analyses Across Timepoints**

| **Model** | **df** | **AIC** | **BIC** | **Χ²** | **RMSEA** | **Χ² diff.** | **RMSEA diff.** | **df diff.** | ***p*** |
| --- | --- | --- | --- | --- | --- | --- | --- | --- | --- |
| Configural | 392 | 15,598.28 | 16,494.35 | 998.22 | 0.115 |  |  |  |  |
| Metric | 428 | 15,564.86 | 16,311.59 | 1,036.80 | 0.110 | 38.58 | -0.005 | 36 | 0.354 |
| Scalar | 464 | 15,519.54 | 16,116.92 | 1,063.48 | 0.105 | 26.68 | -0.005 | 36 | 0.871 |
| Residual | 512 | 15,564.04 | 15,962.29 | 1,203.98 | 0.107 | 140.50 | 0.002 | 48 | <0.001*** |
| **p* < 0.05, ***p* < 0.01, ****p* < 0.001. Confirmatory factor analyses conducted separately at each follow-up timepoint yielded the following fit indices: 3 months, χ²(98) = 231.03, p < .001, CFI = 0.88, TLI = 0.85, RMSEA = 0.115; 6 months, χ²(98) = 222.92, p < .001, CFI = 0.83, TLI = 0.79, RMSEA = 0.131; and 12 months, χ²(98) = 238.23, p < .001, CFI = 0.81, TLI = 0.76, RMSEA = 0.157. | | | | | | | | | |

**Supplementary Table 5. Linear regression models for differences between groups at Cups task performance at baseline without adjusting for clinical and sociodemographic covariates**

| **Proportion of risky choices** | | | | | | | | | | | | |
| --- | --- | --- | --- | --- | --- | --- | --- | --- | --- | --- | --- | --- |
| **Condition** | **Neutral (N = 199)** | | | | **Stress (N = 182)** | | | | **Difference (Stress − Neutral) (N = 179)** | | | |
|  | β(SE) | *Raw p* | Tukey p | Cohen’s d | β(SE) | *Raw p* | Tukey p | Cohen’s d | β(SE) | *Raw p* | Tukey p | Cohen’s d |
| PC vs HC | -0.008 (0.038) | 0.829 | 0.996 | 0.043 | -0.072 (0.046) | 0.116 | 0.394 | 0.330 | -0.028 (0.026) | 0.288 | 0.711 | 0.225 |
| SI vs HC | -0.015 (0.040) | 0.717 | 0.984 | 0.075 | -0.067 (0.046) | 0.150 | 0.472 | 0.305 | -0.057 (0.027) | **0.035*** | 0.147 | 0.455 |
| SA vs HC | 0.024 (0.044) | 0.582 | 0.946 | 0.124 | 0.022 (0.050) | 0.659 | 0.971 | 0.100 | 0.013 (0.029) | 0.650 | 0.969 | 0.104 |
| IQ | -0.003 (0.015) | 0.849 | — | 0.015 | -0.012 (0.017) | 0.497 | — | 0.055 | -0.006 (0.010) | 0.552 | — | 0.048 |
| **EV Sensitivity** | | | | | | | | | | | | |
| **Condition** | **Neutral (N = 199)** | | | | **Stress (N = 182)** | | | | **Difference (Stress − Neutral) (N = 179)** | | | |
|  | β(SE) | *Raw p* | Tukey p | Cohen’s d | β(SE) | *Raw p* | Tukey p | Cohen’s d | β(SE) | *Raw p* | Tukey p | Cohen’s d |
| PC vs HC | 0.078 (0.049) | 0.112 | 0.382 | 0.316 | 0.100 (0.055) | 0.068 | 0.260 | 0.384 | 0.048 (0.046) | 0.308 | 0.737 | 0.215 |
| SI vs HC | 0.077 (0.051) | 0.136 | 0.441 | 0.311 | 0.116 (0.055) | 0.036* | 0.154 | 0.445 | 0.034 (0.047) | 0.466 | 0.885 | 0.156 |
| SA vs HC | 0.104 (0.056) | 0.063 | 0.244 | 0.421 | 0.113 (0.059) | 0.058 | 0.228 | 0.434 | 0.029 (0.051) | 0.566 | 0.939 | 0.132 |
| IQ | -0.118 (0.019) | **<0.001***** | — | 0.476 | -0.114 (0.021) | **<0.001***** | — | 0.436 | 0.011 (0.018) | 0.519 | — | 0.052 |
| *p < 0.05, **p < 0.01, ***p < 0.001. SA = suicide attempt group; SI = suicidal ideation group; PC = psychiatric control group; HC = healthy control group; IQ = Intelligence Quotient, EV – Expected Value. | | | | | | | | | | | | |

**Supplementary Table 6. Linear regression models for differences between groups at Cups task performance at baseline while adjusting for clinical and sociodemographic covariates**

| **Proportion of risky choices** | | | | | | | | | | | | |
| --- | --- | --- | --- | --- | --- | --- | --- | --- | --- | --- | --- | --- |
| **Condition** | **Neutral (N = 199)** | | | | **Stress (N = 182)** | | | | **Difference (Stress − Neutral) (N = 179)** | | | |
|  | β(SE) | *p* | Tukey p | Cohen’s d | β(SE) | *p* | Tukey p | Cohen’s d | β(SE) | *p* | Tukey p | Cohen’s d |
| PC vs HC | -0.008(0.038) | 0.829 | 0.996 | 0.043 | -0.103(0.049) | **0.040*** | 0.166 | 0.473 | -0.028(0.026) | 0.288 | 0.711 | 0.225 |
| SI vs HC | -0.015(0.040) | 0.717 | 0.984 | 0.075 | -0.120(0.054) | **0.028*** | 0.123 | 0.552 | -0.057(0.027) | **0.035*** | 0.147 | 0.455 |
| SA vs HC | 0.024(0.044) | 0.582 | 0.946 | 0.124 | -0.048(0.061) | 0.434 | 0.861 | 0.221 | 0.013(0.029) | 0.650 | 0.969 | 0.104 |
| IQ | -0.003(0.015) | 0.849 |  | 0.015 | -0.011(0.018) | 0.559 |  | 0.049 | -0.006(0.010) | 0.552 |  | 0.048 |
| Social Support (MSPSS) | — | — |  | — | -0.032(0.018) | 0.070 |  | 0.148 | — | — |  | — |
| **EV Sensitivity** | | | | | | | | | | | | |
| **Condition** | **Neutral (N = 199)** | | | | **Stress (N = 182)** | | | | **Difference (Stress − Neutral) (N = 179)** | | | |
|  | **β(SE)** | ***p*** | Tukey p | **Cohen’s d** | **β(SE)** | ***p*** | Tukey p | **Cohen’s d** | **β(SE)** | ***p*** | Tukey p | **Cohen’s d** |
| PC vs HC | 0.044(0.047) | 0.345 | 0.780 | 0.190 | 0.088(0.054) | 0.104 | 0.361 | 0.343 | 0.015(0.048) | 0.756 | 0.989 | 0.073 |
| SI vs HC | 0.045(0.051) | 0.369 | 0.805 | 0.194 | 0.120(0.054) | **0.028*** | 0.122 | 0.469 | 0.028(0.059) | 0.630 | 0.963 | 0.140 |
| SA vs HC | 0.047(0.055) | 0.388 | 0.823 | 0.202 | 0.108(0.058) | 0.063 | 0.245 | 0.425 | 0.108(0.063) | 0.089 | 0.322 | 0.531 |
| IQ | -0.122(0.019) | **<0.001*** |  | 0.520 | -0.123(0.021) | **<0.001*** |  | 0.484 | -0.019(0.022) | 0.372 |  | 0.095 |
| Female | 0.107(0.035) | **0.003**** |  | 0.457 | 0.119(0.039) | **0.003**** |  | 0.467 | — | — |  | — |
| White | -0.100(0.039) | **0.011*** |  | 0.429 | — | — |  | — | — | — |  | — |
| CTQ | 0.004(0.002) | **0.034*** |  | 0.017 | — | — |  | — | -0.006(0.002) | **0.004**** |  | 0.028 |
| SES | — | — |  | — | — | — |  | — | 0.085(0.025) | **<0.001*** |  | 0.420 |
| Factor 2- Anhedonia | — | — |  | — | — | — |  | — | 0.059(0.023) | **0.011*** |  | 0.290 |
| *p < 0.05, **p < 0.01, ***p < 0.001. SA = suicide attempt group; SI = suicidal ideation group; PC = psychiatric control group; HC = healthy control group; IQ = Intelligence Quotient, EV – Expected Value. | | | | | | | | | | | | |

**Supplementary Table 7. Linear mixed effects models for differences between groups at changes in Cups task performance over time without adjusting for clinical and sociodemographic covariates IPW weighted analysis**

| **Proportion of risky choices** | | | | | | | | | | | | |
| --- | --- | --- | --- | --- | --- | --- | --- | --- | --- | --- | --- | --- |
| **Condition** | **Neutral (N = 202)** | | | | **Stress (N = 192)** | | | | **Difference (Stress − Neutral) (N = 189)** | | | |
|  | β(SE) | *p* | Tukey p | Cohen’s d | β(SE) | *p* | Tukey p | Cohen’s d | β(SE) | *p* | Tukey p | Cohen’s d |
| PC vs HC | -0.020(0.040) | 0.616 | 0.958 | 0.101 | -0.055(0.046) | 0.228 | 0.621 | 0.249 | -0.027(0.028) | 0.334 | 0.767 | 0.204 |
| SI vs HC | -0.023(0.041) | 0.577 | 0.944 | 0.116 | -0.074(0.047) | 0.112 | 0.383 | 0.335 | -0.058(0.028) | **0.037*** | **0.155** | 0.446 |
| SA vs HC | 0.007(0.043) | 0.875 | 0.999 | 0.035 | 0.036(0.049) | 0.471 | 0.888 | 0.161 | 0.022(0.029) | 0.457 | 0.878 | 0.166 |
| Time | 0.015(0.012) | 0.232 |  | 0.076 | 0.017(0.013) | 0.182 |  | 0.078 | 0.002(0.010) | 0.873 |  | 0.012 |
| PC × Time | 0.018(0.020) | 0.377 | 0.810 | 0.090 | -0.005(0.022) | 0.829 | 0.996 | 0.021 | -0.026(0.017) | 0.119 | 0.402 | 0.201 |
| SI × Time | 0.021(0.021) | 0.321 | 0.751 | 0.108 | 0.015(0.021) | 0.478 | 0.890 | 0.068 | 0.005(0.017) | 0.746 | 0.988 | 0.041 |
| SA × Time | -0.024(0.017) | 0.167 | 0.504 | 0.121 | -0.119(0.021) | **<0.001***** | **<0.001***** | 0.534 | -0.045(0.017) | **0.007**** | **0.037*** | 0.346 |
| IQ | -0.010(0.013) | 0.469 |  | 0.049 | -0.006(0.016) | 0.707 |  | 0.027 | -0.002(0.009) | 0.843 |  | 0.013 |
| **EV Sensitivity** | | | | | | | | | | | | |
| **Condition** | **Neutral (N = 202)** | | | | **Stress (N = 192)** | | | | **Difference (Stress − Neutral) (N = 189)** | | | |
|  | β(SE) | *p* | Tukey p | Cohen’s d | β(SE) | *p* | Tukey p | Cohen’s d | β(SE) | *p* | Tukey p | Cohen’s d |
| PC vs HC | 0.052(0.053) | 0.326 | 0.757 | 0.199 | 0.093(0.059) | 0.113 | 0.385 | 0.331 | 0.056(0.048) | 0.251 | 0.657 | 0.243 |
| SI vs HC | 0.058(0.054) | 0.285 | 0.707 | 0.222 | 0.098(0.059) | 0.097 | 0.342 | 0.350 | 0.027(0.049) | 0.575 | 0.943 | 0.119 |
| SA vs HC | 0.077(0.057) | 0.177 | 0.528 | 0.296 | 0.102(0.062) | 0.101 | 0.352 | 0.364 | 0.032(0.051) | 0.529 | 0.922 | 0.139 |
| Time | -0.003(0.018) | 0.846 |  | 0.013 | 0.007(0.020) | 0.746 |  | 0.023 | 0.012(0.019) | 0.507 |  | 0.054 |
| PC × Time | 0.018(0.029) | 0.539 | 0.926 | 0.068 | -0.014(0.034) | 0.676 | 0.975 | 0.051 | -0.035(0.031) | 0.258 | 0.668 | 0.153 |
| SI × Time | 0.035(0.031) | 0.253 | 0.660 | 0.135 | 0.027(0.033) | 0.409 | 0.839 | 0.098 | -0.009(0.031) | 0.764 | 0.990 | 0.041 |
| SA × Time | -0.001(0.024) | 0.981 | 1.000 | 0.002 | -0.029(0.034) | 0.384 | 0.817 | 0.104 | -0.023(0.031) | 0.453 | 0.875 | 0.101 |
| IQ | -0.122(0.017) | **<0.001*** |  | 0.469 | -0.117(0.019) | **<0.001*** |  | 0.417 | 0.005(0.015) | 0.723 |  | 0.024 |
| *p < 0.05, **p < 0.01, ***p < 0.001. SA = suicide attempt group; SI = suicidal ideation group; PC = psychiatric control group; HC = healthy control group; IQ = Intelligence Quotient, EV – Expected Value. | | | | | | | | | | | | |

**Supplementary Table 8. Sensitivity analysis- Linear mixed effects models for differences between groups at changes in Cups task performance over time while adjusting for clinical and sociodemographic covariates non weighted analysis**

| **Proportion of risky choices** | | | | | | | | | | | | |
| --- | --- | --- | --- | --- | --- | --- | --- | --- | --- | --- | --- | --- |
| **Condition** | **Neutral (N = 202)** | | | | **Stress (N = 192)** | | | | **Difference (Stress − Neutral) (N = 189)** | | | |
|  | β(SE) | *p* | Tukey p | Cohen’s d | β(SE) | *p* | Tukey p | Cohen’s d | β(SE) | *p* | Tukey p | Cohen’s d |
| PC vs HC | 0.004(0.035) | 0.899 | 0.999 | 0.024 | -0.042(0.042) | 0.318 | 0.749 | 0.195 | -0.014(0.025) | 0.575 | 0.944 | 0.117 |
| SI vs HC | -0.006(0.037) | 0.873 | 0.999 | 0.031 | -0.059(0.043) | 0.175 | 0.526 | 0.273 | -0.082(0.029) | 0.005 | 0.024 | 0.685 |
| SA vs HC | 0.034(0.040) | 0.398 | 0.832 | 0.180 | 0.032(0.047) | 0.491 | 0.901 | 0.150 | -0.021(0.031) | 0.486 | 0.898 | 0.179 |
| Time | 0.016(0.010) | 0.112 | — | 0.084 | 0.020(0.012) | 0.091 | — | 0.092 | 0.005(0.009) | 0.606 | — | 0.040 |
| PC × Time | 0.006(0.017) | 0.699 | 0.980 | 0.034 | -0.015(0.021) | 0.482 | 0.897 | 0.070 | -0.023(0.015) | 0.130 | 0.432 | 0.195 |
| SI × Time | 0.009(0.019) | 0.651 | 0.969 | 0.045 | 0.004(0.021) | 0.837 | 0.997 | 0.020 | 0.002(0.016) | 0.890 | 0.999 | 0.018 |
| SA × Time | -0.056(0.020) | **0.006**** | **0.031*** | 0.294 | -0.092(0.025) | **<0.001***** | **0.002**** | 0.427 | -0.024(0.019) | 0.205 | 0.588 | 0.202 |
| IQ | 0.004(0.013) | 0.762 | — | 0.021 | -0.002(0.015) | 0.909 | — | 0.008 | -0.008(0.009) | 0.366 | — | 0.067 |
| Female | -0.065(0.025) | 0.010 | — | 0.343 | — | — | — | — | — | — | — | — |
| Factor 2- Anhedonia | — | — | — | — | — | — | — | — | 0.024(0.011) | 0.032 | — | 0.204 |
| **EV Sensitivity** | | | | | | | | | | | | |
| **Condition** | **Neutral (N = 202)** | | | | **Stress (N = 192)** | | | | **Difference (Stress − Neutral) (N = 189)** | | | |
|  | β(SE) | *p* | Tukey p | Cohen’s d | β(SE) | *p* | Tukey p | Cohen’s d | β(SE) | *p* | Tukey p | Cohen’s d |
| PC vs HC | 0.038 (0.046) | 0.405 | 0.838 | 0.247 | 0.094 (0.054) | 0.081 | 0.300 | 0.525 | 0.030 (0.040) | 0.455 | 0.877 | 0.156 |
| SI vs HC | 0.051 (0.048) | 0.289 | 0.713 | 0.331 | 0.114 (0.055) | 0.038 | 0.162 | 0.635 | 0.007 (0.048) | 0.876 | 0.999 | 0.038 |
| SA vs HC | 0.062 (0.053) | 0.244 | 0.647 | 0.399 | 0.088 (0.060) | 0.140 | 0.451 | 0.495 | 0.053 (0.051) | 0.295 | 0.721 | 0.275 |
| Time | −0.007 (0.013) | 0.608 | — | 0.027 | 0.004 (0.016) | 0.798 | — | 0.015 | 0.006 (0.016) | 0.691 | — | 0.032 |
| PC × Time | 0.042 (0.022) | 0.057 | 0.213 | 0.170 | −0.006 (0.029) | 0.825 | 0.996 | 0.023 | −0.040 (0.026) | 0.129 | 0.432 | 0.205 |
| SI × Time | 0.009 (0.025) | 0.722 | 0.984 | 0.036 | −0.009 (0.029) | 0.746 | 0.989 | 0.034 | 0.006 (0.027) | 0.812 | 0.995 | 0.033 |
| SA × Time | −0.006 (0.026) | 0.825 | 0.996 | 0.024 | −0.034 (0.034) | 0.321 | 0.755 | 0.124 | −0.004 (0.033) | 0.899 | 0.999 | 0.022 |
| IQ | −0.117 (0.017) | **<0.001***** | — | 0.474 | −0.117 (0.019) | **<0.001***** | — | 0.429 | −0.006 (0.014) | 0.681 | — | 0.029 |
| Female | 0.103 (0.033) | **0.002**** | — | 0.416 | 0.127 (0.037) | **<0.001***** | — | 0.468 | — | — | — | — |
| White | −0.114 (0.035) | **0.001**** | — | 0.461 | −0.088 (0.041) | **0.034*** | — | 0.324 | — | — | — | — |
| SES | — | — | — | — | — | — | — | — | 0.059 (0.017) | <0.001 | — | 0.307 |
| Factor 2- Anhedonia | — | — | — | — | — | — | — | — | 0.049 (0.019) | 0.009 | — | 0.251 |
| *p < 0.05, **p < 0.01, ***p < 0.001. SA = suicide attempt group; SI = suicidal ideation group; PC = psychiatric control group; HC = healthy control group; IQ = Intelligence Quotient, EV – Expected Value. | | | | | | | | | | | | |

**Supplementary Table 9. Person correlations between biological measures and Cups task performance at baseline adjusting for FDR using Benjamini-Yekuteali method**

| **Proportion of risky choices** | | | | | | | | | |
| --- | --- | --- | --- | --- | --- | --- | --- | --- | --- |
| **Condition** | **Neutral** | | | **Stress** | | | **Difference (Stress − Neutral)** | | |
|  | **Pearson r** | **p** | **Adjusted p** | **Pearson r** | **p** | **Adjusted p** | **Pearson r** | **p** | **Adjusted p** |
| CRP | -0.125 | 0.086 | 0.525 | -0.123 | 0.105 | 1.000 | -0.040 | 0.602 | 1.000 |
| HCC | 0.073 | 0.351 | 1.000 | -0.020 | 0.804 | 1.000 | -0.137 | 0.091 | 1.000 |
| IL10 | -0.035 | 0.625 | 1.000 | 0.009 | 0.905 | 1.000 | 0.048 | 0.526 | 1.000 |
| IL1ra | -0.115 | 0.107 | 0.525 | -0.055 | 0.471 | 1.000 | 0.011 | 0.884 | 1.000 |
| IL6 | -0.155 | 0.029* | 0.430 | -0.068 | 0.367 | 1.000 | 0.073 | 0.336 | 1.000 |
| TNF-α | -0.068 | 0.343 | 1.000 | 0.001 | 0.987 | 1.000 | 0.088 | 0.248 | 1.000 |
| **EV Sensitivity** | | | | | | | | | |
| **Condition** | **Neutral** | | | **Stress** | | | **Difference (Stress − Neutral)** | | |
|  | **Pearson r** | **p** | **Adjusted p** | **Pearson r** | **p** | **Adjusted p** | **Pearson r** | **p** | **Adjusted p** |
| CRP | 0.198 | 0.006** | **0.044*** | 0.203 | 0.007** | 0.052 | 0.025 | 0.740 | 1.000 |
| HCC | 0.083 | 0.289 | 0.745 | -0.028 | 0.728 | 1.000 | -0.121 | 0.139 | 1.000 |
| IL10 | 0.104 | 0.144 | 0.529 | 0.121 | 0.107 | 0.526 | 0.000 | 1.000 | 1.000 |
| IL1ra | 0.112 | 0.115 | 0.529 | 0.048 | 0.527 | 1.000 | -0.002 | 0.981 | 1.000 |
| IL6 | 0.242 | <0.001* | **0.009**** | 0.243 | 0.001** | **0.016*** | 0.036 | 0.634 | 1.000 |
| TNF-α | 0.073 | 0.304 | 0.745 | 0.017 | 0.825 | 1.000 | -0.046 | 0.542 | 1.000 |
| Adjusted p present false detection rate (FDR) corrected p- value according to Benjamini-Yukutaeli method. *p < 0.05, **p < 0.01, ***p < 0.001. | | | | | | | | | |

**Supplementary Table 10. Linear regression models for baseline biological measures and Cups task performance at baseline adjusted for sociodemographic and clinical covariates**

| **EV Sensitivity** | | | | | | | | | | | |
| --- | --- | --- | --- | --- | --- | --- | --- | --- | --- | --- | --- |
| **Condition** | **Neutral** | | |  | **Neutral** | | |  | **Stress** | | |
|  | β (SE) | p | Cohen’s d |  | β (SE) | p | Cohen’s d |  | β (SE) | p | Cohen’s d |
| CRP | −0.018 (0.074) | 0.806 | 0.082 | IL-6 | 0.070 (0.036) | 0.055 | 0.318 | IL-6 | 0.078 (0.041) | 0.060 | 0.297 |
| PC vs HC | 0.018 (0.053) | 0.732 | 0.081 | PC vs HC | 0.022 (0.052) | 0.676 | 0.098 | PC vs HC | 0.080 (0.059) | 0.176 | 0.303 |
| SI vs HC | 0.057 (0.057) | 0.315 | 0.254 | SI vs HC | 0.054 (0.055) | 0.328 | 0.246 | SI vs HC | 0.144 (0.058) | **0.015*** | 0.548 |
| SA vs HC | −0.021 (0.073) | 0.770 | 0.095 | SA vs HC | −0.006 (0.070) | 0.933 | 0.027 | SA vs HC | 0.112 (0.068) | 0.102 | 0.428 |
| IQ | −0.062 (0.025) | **0.012*** | 0.278 | IQ | −0.061 (0.023) | **0.009**** | 0.278 | IQ | -0.117 (0.024) | **<0.001***** | 0.445 |
| BMI | 0.006 (0.003) | 0.050 | 0.029 | BMI | 0.003 (0.003) | 0.314 | 0.014 | BMI | 0.001 (0.026) | 0.985 | 0.002 |
| Female | 0.136 (0.041) | **0.001**** | 0.604 | Female | 0.122 (0.040) | **0.002**** | 0.554 | Female | 0.125 (0.044) | **0.006**** | 0.477 |
| Factor 3- Affective Lability | 0.047 (0.024) | 0.052 | 0.208 | Factor 3 | 0.045 (0.023) | 0.052 | 0.202 |  |  |  |  |
| CTQ | 0.007 (0.002) | **0.002**** | 0.033 | CTQ | 0.007 (0.002) | **0.002**** | 0.032 |  |  |  |  |
| EV sensitivity- Expected Value Sensitivity . *p < 0.05, **p < 0.01, ***p < 0.001. | | | | | | | | | | | |

**Supplementary Table 11. Linear mixed effects models for the association between changes in biomarkers and changes in Cups task performance over time without adjusting for clinical and sociodemographic covariates IPW weighted analysis**

|  | **Proportion of risky choices** | | | | | | | | |
| --- | --- | --- | --- | --- | --- | --- | --- | --- | --- |
| **Condition** | **Neutral** | | | **Stress** | | | **Difference (Stress − Neutral)** | | |
|  | **β(SE)** | ***p*** | **Adjusted p** | **β(SE)** | ***p*** | **Adjusted p** | **β(SE)** | ***p*** | **Adjusted p** |
| **CRP** | -0.115(0.025) | **<0.001*** | **<0.001*** | -0.111(0.029) | **<0.001*** | **0.009**** | -0.003(0.020) | 0.880 | 1.000 |
| Time | 0.010(0.010) | 0.314 | 1.000 | 0.003(0.011) | 0.781 | 1.000 | -0.004(0.008) | 0.601 | 1.000 |
| CRP × Time | -0.000(0.021) | 0.988 | 1.000 | -0.047(0.026) | 0.074 | 0.744 | -0.034(0.019) | 0.070 | 0.744 |
| **HCC** | 0.005(0.016) | 0.756 | 1.000 | -0.001(0.019) | 0.974 | 1.000 | -0.003(0.012) | 0.818 | 1.000 |
| Time | -0.019(0.033) | 0.567 | 1.000 | -0.067(0.038) | 0.084 | 1.000 | -0.019(0.025) | 0.442 | 1.000 |
| HCC × Time | 0.017(0.015) | 0.275 | 1.000 | 0.032(0.017) | 0.066 | 0.744 | 0.004(0.011) | 0.720 | 1.000 |
| **IL10** | -0.076(0.031) | **0.014*** | 0.419 | -0.046(0.036) | 0.199 | 1.000 | 0.035(0.022) | 0.109 | 1.000 |
| Time | 0.002(0.025) | 0.930 | 1.000 | 0.069(0.030) | **0.022*** | 0.827 | 0.070(0.020) | **<0.001*** | 1.000 |
| IL10 × Time | 0.009(0.021) | 0.681 | 1.000 | -0.062(0.024) | **0.010*** | 0.366 | -0.070(0.016) | **<0.001*** | **0.003**** |
| **IL1ra** | -0.030(0.015) | **0.040*** | 0.850 | -0.025(0.018) | 0.161 | 1.000 | -0.007(0.011) | 0.548 | 1.000 |
| Time | -0.001(0.053) | 0.984 | 1.000 | 0.057(0.067) | 0.392 | 1.000 | 0.029(0.045) | 0.517 | 1.000 |
| IL1ra × Time | 0.002(0.009) | 0.836 | 1.000 | -0.012(0.012) | 0.334 | 1.000 | -0.008(0.008) | 0.348 | 1.000 |
| **IL6** | -0.073(0.016) | **<0.001*** | **<0.001*** | -0.060(0.019) | **0.002**** | 0.076 | 0.010(0.013) | 0.417 | 1.000 |
| Time | 0.003(0.015) | 0.843 | 1.000 | 0.021(0.017) | 0.223 | 1.000 | 0.017(0.012) | 0.152 | 1.000 |
| IL6 × Time | 0.008(0.012) | 0.518 | 1.000 | -0.026(0.014) | 0.065 | 0.744 | -0.029(0.010) | **0.004**** | 0.197 |
| **TNF-α** | -0.013(0.006) | **0.029*** | 0.723 | -0.010(0.007) | 0.167 | 1.000 | 0.004(0.004) | 0.381 | 1.000 |
| Time | 0.049(0.035) | 0.162 | 1.000 | 0.075(0.042) | 0.075 | 1.000 | 0.019(0.028) | 0.495 | 1.000 |
| TNF-α × Time | -0.006(0.005) | 0.242 | 1.000 | -0.012(0.006) | **0.045*** | 0.744 | -0.005(0.004) | 0.258 | 1.000 |
|  | **EV Sensitivity** | | | | | | | | |
| **Condition** | **Neutral** | | | **Stress** | | | **Difference (Stress − Neutral)** | | |
|  | **β(SE)** | ***p*** | **Adjusted p** | **β(SE)** | ***p*** | **Adjusted p** | **β(SE)** | ***p*** | **Adjusted p** |
| **CRP** | -0.070(0.036) | 0.055 | 1.000 | -0.065(0.041) | 0.116 | 1.000 | 0.010(0.035) | 0.764 | 1.000 |
| Time | -0.012(0.014) | 0.371 | 1.000 | -0.038(0.016) | **0.019*** | 0.827 | -0.023(0.014) | 0.121 | 1.000 |
| CRP × Time | 0.071(0.029) | **0.014*** | 0.366 | 0.118(0.036) | **0.002**** | 0.121 | 0.083(0.034) | **0.015*** | 0.366 |
| **HCC** | 0.014(0.024) | 0.554 | 1.000 | 0.018(0.026) | 0.494 | 1.000 | -0.004(0.020) | 0.828 | 1.000 |
| Time | 0.067(0.048) | 0.162 | 1.000 | 0.010(0.056) | 0.860 | 1.000 | -0.020(0.047) | 0.671 | 1.000 |
| HCC × Time | -0.034(0.022) | 0.124 | 1.000 | -0.012(0.025) | 0.643 | 1.000 | 0.005(0.021) | 0.795 | 1.000 |
| **IL10** | 0.021(0.046) | 0.648 | 1.000 | 0.032(0.051) | 0.531 | 1.000 | 0.043(0.040) | 0.279 | 1.000 |
| Time | 0.015(0.036) | 0.670 | 1.000 | 0.040(0.043) | 0.351 | 1.000 | 0.069(0.038) | 0.070 | 1.000 |
| IL10 × Time | -0.005(0.030) | 0.875 | 1.000 | -0.041(0.035) | 0.240 | 1.000 | -0.061(0.031) | **0.048*** | 0.744 |
| **IL1ra** | 0.024(0.021) | 0.260 | 1.000 | 0.010(0.025) | 0.671 | 1.000 | -0.004(0.019) | 0.817 | 1.000 |
| Time | -0.029(0.074) | 0.697 | 1.000 | -0.068(0.093) | 0.466 | 1.000 | -0.151(0.081) | 0.064 | 1.000 |
| IL1ra × Time | 0.007(0.013) | 0.584 | 1.000 | 0.011(0.017) | 0.510 | 1.000 | 0.027(0.015) | 0.066 | 0.744 |
| **IL6** | -0.014(0.025) | 0.582 | 1.000 | -0.011(0.028) | 0.702 | 1.000 | 0.004(0.023) | 0.844 | 1.000 |
| Time | -0.024(0.021) | 0.261 | 1.000 | -0.058(0.025) | **0.021*** | 0.827 | -0.034(0.022) | 0.122 | 1.000 |
| IL6 × Time | 0.032(0.017) | 0.070 | 0.744 | 0.048(0.020) | **0.020*** | 0.437 | 0.031(0.018) | 0.088 | 0.829 |
| **TNF-α** | -0.001(0.009) | 0.868 | 1.000 | -0.009(0.010) | 0.384 | 1.000 | -0.003(0.008) | 0.711 | 1.000 |
| Time | 0.040(0.051) | 0.425 | 1.000 | -0.038(0.060) | 0.528 | 1.000 | -0.041(0.052) | 0.423 | 1.000 |
| TNF-α × Time | -0.005(0.008) | 0.536 | 1.000 | 0.004(0.009) | 0.615 | 1.000 | 0.006(0.008) | 0.443 | 1.000 |
| Adjusted p present false detection rate (FDR) corrected p- value according to Benjamini-Yukutaeli method *p < 0.05, **p < 0.01, ***p < 0.001. IQ = Intelligence Quotient, EV – Expected Value. | | | | | | | | | |

**Supplementary Table 12. Sensitivity analysis- non-IPW weighted linear mixed effects models for the association between changes in biomarkers and changes in Cups task performance over time while adjusting for clinical and sociodemographic covariates**

| **Proportion of risky choices** | | | | | | | |
| --- | --- | --- | --- | --- | --- | --- | --- |
| **Condition** | **Neutral (N=177)** | | |  | **Neutral (N=165)** | | |
|  | β (SE) | p | Cohen’s d |  | β (SE) | p | Cohen’s d |
| CRP | −0.120 (0.043) | **0.006**** | 0.653 | IL-6 | −0.062 (0.023) | **0.007**** | 0.342 |
| Time | 0.006 (0.009) | 0.485 | 0.035 | Time | 0.002 (0.015) | 0.892 | 0.011 |
| CRP × Time | 0.027 (0.023) | 0.247 | 0.147 | IL-6 × Time | 0.011 (0.013) | 0.387 | 0.061 |
| PC vs HC | 0.000 (0.033) | 0.992 | 0.002 | PC vs HC | 0.020 (0.033) | 0.548 | 0.109 |
| SI vs HC | −0.011 (0.036) | 0.770 | 0.057 | SI vs HC | −0.012 (0.035) | 0.732 | 0.067 |
| SA vs HC | 0.010 (0.042) | 0.820 | 0.052 | SA vs HC | 0.015 (0.041) | 0.706 | 0.085 |
| IQ | 0.005 (0.014) | 0.696 | 0.029 | IQ | 0.011 (0.013) | 0.414 | 0.060 |
| BMI | 0.009 (0.015) | 0.545 | 0.051 | BMI | 0.018 (0.015) | 0.224 | 0.101 |
|  |  |  |  | Female | −0.068 (0.026) | **0.009**** | 0.374 |
| **Condition** | **Stress (N=163)** | | |  | **Difference (Stress − Neutral) (N=150)** | | |
|  | β (SE) | p | Cohen’s d |  | β (SE) | p | Cohen’s d |
| CRP | −0.136 (0.048) | **0.006**** | 0.650 | IL-10 | 0.015 (0.030) | 0.605 | 0.129 |
| Time | 0.003 (0.010) | 0.772 | 0.014 | Time | 0.021 (0.025) | 0.399 | 0.175 |
| CRP × Time | 0.012 (0.027) | 0.661 | 0.057 | IL-10 × Time | −0.023 (0.021) | 0.277 | 0.195 |
| PC vs HC | −0.043 (0.040) | 0.281 | 0.206 | PC vs HC | −0.027 (0.022) | 0.222 | 0.230 |
| SI vs HC | −0.046 (0.042) | 0.266 | 0.223 | SI vs HC | −0.083 (0.027) | **0.003**** | 0.696 |
| SA vs HC | 0.013 (0.049) | 0.787 | 0.064 | SA vs HC | −0.058 (0.032) | 0.078 | 0.482 |
| IQ | 0.001 (0.016) | 0.974 | 0.003 | IQ | −0.007 (0.010) | 0.451 | 0.062 |
| BMI | 0.026 (0.019) | 0.154 | 0.127 | BMI | −0.001 (0.009) | 0.944 | 0.005 |
|  |  |  |  | Factor 2- Anhedonia | 0.035 (0.012) | **0.005**** | 0.294 |
| *p < 0.05, **p < 0.01, ***p < 0.001. | | | | | | | |

**Supplementary Table 13. Logistic models for Cups performance predicting SA and SB outcomes unadjusted for clinical covariates**

| **SA outcome (Number of events = 15)** | | | | | | | | | | | |
| --- | --- | --- | --- | --- | --- | --- | --- | --- | --- | --- | --- |
| **Proportion of risky choices** | | | | | | | | | | | |
| **Neutral** | | | | **Stress** | | | | **Difference (Stress − Neutral)** | | | |
| OR (SE) | 95% CI | p | Cohen’s d | OR (SE) | 95% CI | p | Cohen’s d | OR (SE) | 95% CI | p | Cohen’s d |
| 1.487 (0.284) | 0.853–2.593 | 0.162 | 0.219 | 1.467 (0.292) | 0.828–2.597 | 0.189 | 0.211 | 1.083 (0.277) | 0.629–1.863 | 0.775 | 0.044 |
| **EV Sensitivity** | | | | | | | | | | | |
| **Neutral** | | | | **Stress** | | | | **Difference (Stress − Neutral)** | | | |
| OR (SE) | 95% CI | p | Cohen’s d | OR (SE) | 95% CI | p | Cohen’s d | OR (SE) | 95% CI | p | Cohen’s d |
| 0.950 (0.273) | 0.556–1.624 | 0.852 | 0.028 | 0.631 (0.287) | 0.360–1.107 | 0.109 | 0.254 | 0.609 (0.281) | 0.351–1.056 | 0.077 | 0.273 |
| **SB outcome (Number of events = 32)** | | | | | | | | | | | |
| **Proportion of risky choices** | | | | | | | | | | | |
| **Neutral** | | | | **Stress** | | | | **Difference (Stress − Neutral)** | | | |
| OR (SE) | 95% CI | p | Cohen’s d | OR (SE) | 95% CI | p | Cohen’s d | OR (SE) | 95% CI | p | Cohen’s d |
| 0.939 (0.198) | 0.637–1.385 | 0.752 | 0.035 | 0.922 (0.204) | 0.618–1.376 | 0.690 | 0.045 | 0.881 (0.206) | 0.588–1.319 | 0.537 | 0.070 |
| **EV Sensitivity** | | | | | | | | | | | |
| **Neutral** | | | | **Stress** | | | | **Difference (Stress − Neutral)** | | | |
| OR (SE) | 95% CI | p | Cohen’s d | OR (SE) | 95% CI | p | Cohen’s d | OR (SE) | 95% CI | p | Cohen’s d |
| 1.118 (0.200) | 0.756–1.654 | 0.577 | 0.061 | 0.700 (0.208) | 0.466–1.054 | 0.087 | 0.196 | 0.553 (0.223) | 0.358–0.856 | **0.008**** | 0.326 |
| EV sensitivity- Expected Value Sensitivity . *p < 0.05, **p < 0.01, ***p < 0.001. | | | | | | | | | | | |

**Supplementary Table 14. Cox models for Cups performance predicting time to onset of SA and SB outcomes unadjusted for clinical covariates**

| **SA outcome (Number of events = 15)** | | | | | | | | |
| --- | --- | --- | --- | --- | --- | --- | --- | --- |
| **Proportion of risky choices** | | | | | | | | |
| **Neutral** | | | **Stress** | | | **Difference (Stress − Neutral)** | | |
| HR | 95% CI | p | HR | 95% CI | p | HR | 95% CI | p |
| 1.281 | 0.762–2.151 | 0.350 | 1.251 | 0.840–1.862 | 0.271 | 1.019 | 0.684–1.517 | 0.927 |
| **EV Sensitivity** | | | | | | | | |
| **Neutral** | | | **Stress** | | | **Difference (Stress − Neutral)** | | |
| HR | 95% CI | p | HR | 95% CI | p | HR | 95% CI | p |
| 1.130 | 0.751–1.700 | 0.558 | 0.839 | 0.576–1.220 | 0.357 | 0.619 | 0.401–0.956 | **0.031*** |
| **SB outcome (Number of events = 32)** | | | | | | | | |
| **Proportion of risky choices** | | | | | | | | |
| **Neutral** | | | **Stress** | | | **Difference (Stress − Neutral)** | | |
| HR | 95% CI | p | HR | 95% CI | p | HR | 95% CI | p |
| 0.843 | 0.555–1.280 | 0.423 | 0.881 | 0.604–1.284 | 0.509 | 0.922 | 0.684–1.243 | 0.596 |
| **EV Sensitivity** | | | | | | | | |
| **Neutral** | | | **Stress** | | | **Difference (Stress − Neutral)** | | |
| HR | 95% CI | p | HR | 95% CI | p | HR | 95% CI | p |
| 1.011 | 0.557–1.836 | 0.971 | 0.690 | 0.417–1.141 | 0.148 | 0.697 | 0.466–1.043 | 0.080 |
| EV sensitivity- Expected Value Sensitivity . *p < 0.05, **p < 0.01, ***p < 0.001. | | | | | | | | |

**Supplementary Table 15. Cox models for Cups performance predicting time to onset of SA and SB outcomes adjusting for clinical covariates (social support, PTSD and Factor 2)**

|  | **SA outcome (Number of events = 13)** | | | | | | | | |
| --- | --- | --- | --- | --- | --- | --- | --- | --- | --- |
|  | **Proportion of risky choices** | | | | | | | | |
| **Predictor** | **Neutral** | | | **Stress** | | | **Difference (Stress − Neutral)** | | |
|  | HR | 95% CI | p | HR | 95% CI | p | HR | 95% CI | p |
| Risky responses | 1.283 | 0.748–2.200 | 0.365 | 1.157 | 0.754–1.777 | 0.505 | 0.811 | 0.510–1.290 | 0.377 |
| Social support | 0.566 | 0.402–0.797 | 0.001 | 0.537 | 0.389–0.742 | <0.001 | 0.505 | 0.342–0.744 | <0.001 |
|  | **EV Sensitivity** | | | | | | | | |
| **Predictor** | **Neutral** | | | **Stress** | | | **Difference (Stress − Neutral)** | | |
|  | HR | 95% CI | p | HR | 95% CI | p | HR | 95% CI | p |
| EV Sensitivity | 0.948 | 0.477–1.883 | 0.878 | 0.664 | 0.366–1.204 | 0.178 | 0.625 | 0.367–1.063 | 0.083 |
| Social support | 0.560 | 0.405–0.776 | <0.001 | 0.533 | 0.389–0.731 | <0.001 | 0.530 | 0.393–0.715 | <0.001 |
|  | **SB outcome (Number of events = 22)** | | | | | | | | |
|  | **Proportion of risky choices** | | | | | | | | |
| **Predictor** | **Neutral** | | | **Stress** | | | **Difference (Stress − Neutral)** | | |
|  | HR | 95% CI | p | HR | 95% CI | p | HR | 95% CI | p |
| Risky responses | 0.811 | 0.478–1.377 | 0.438 | 0.703 | 0.445–1.112 | 0.132 | 0.798 | 0.569–1.117 | 0.188 |
| Current PTSD | 3.846 | 1.693–8.736 | 0.001 | 3.243 | 1.435–7.327 | 0.005 | 3.620 | 1.532–8.554 | 0.003 |
| Factor 2- Anhedonia | 1.664 | 1.050–2.636 | 0.030 | 1.911 | 1.164–3.135 | 0.010 | 1.761 | 1.082–2.866 | 0.023 |
|  | **EV Sensitivity** | | | | | | | | |
| **Predictor** | **Neutral** | | | **Stress** | | | **Difference (Stress − Neutral)** | | |
|  | HR | 95% CI | p | HR | 95% CI | p | HR | 95% CI | p |
| EV Sensitivity | 1.386 | 0.840–2.285 | 0.201 | 1.041 | 0.692–1.568 | 0.846 | 0.638 | 0.395–1.030 | 0.066 |
| Current PTSD | 4.299 | 1.758–10.511 | 0.001 | 3.158 | 1.399–7.126 | 0.006 | 3.963 | 1.580–9.942 | 0.003 |
| Factor 2- Anhedonia | 1.692 | 1.125–2.544 | 0.011 | 1.837 | 1.138–2.966 | 0.013 | 1.873 | 1.099–3.191 | 0.021 |
|  | EV sensitivity- Expected Value Sensitivity . *p < 0.05, **p < 0.01, ***p < 0.001. | | | | | | | | |
